# Genome-wide meta-analysis of shingles identifies eight susceptibility loci implicating antigen presentation and interferon signaling

**DOI:** 10.64898/2026.08.14.26360428

**Authors:** Hele Haapaniemi, Satu Strausz, Tommi Strausz, Estonian Biobank Research Team, FinnGen, Anssi Lipponen, Ville Leinonen, Mikko Hiltunen, Sami Heikkinen, Erik Abner, Hanna M. Ollila

**Author notes:** Corresponding author: Hanna M. Ollila. Lists of authors and their affiliations appear at the end of the paper (Estonian Biobank Research Team, FinnGen).

## Abstract

Shingles (herpes zoster), caused by reactivation of varicella zoster virus (VZV), affects about one third of people during their lifetime, and host genetics contribute to susceptibility. We performed a genome-wide association study (GWAS) meta-analysis across five cohorts comprising 70,216 cases and 1,646,257 controls of European ancestry. We identified eight genome-wide significant loci, replicating associations in the HLA region and near *IFNA21* and identifying novel associations near *BACH2*, *IFNAR2*, *RHOBTB1*, *KPNB1* and *CPN1* and at the *IGH* locus. HLA fine-mapping identified eight independent allele signals, six at HLA-B, with the strongest protective association for HLA-B*44:02/B*44:27. Colocalization linked the risk alleles to lower expression of *IFNAR2* in T cells, NK cells and skin, *BACH2* in CD4⁺ T cells, and *IGHG1* and *IGHG2* in B cells. These loci implicate MHC class I antigen presentation, type I interferon signaling, T-cell memory and humoral immunity. Shingles was genetically correlated with pain, infectious and immune-mediated diseases. In Mendelian randomization, genetic liability to shingles was associated with multiple sclerosis and systemic lupus erythematosus, but both associations depended on the MHC instrument, suggesting shared HLA effects. Genetic liability to pain was associated with higher shingles risk. These findings advance understanding of the genetic architecture of VZV reactivation.

**Graphical abstract:** 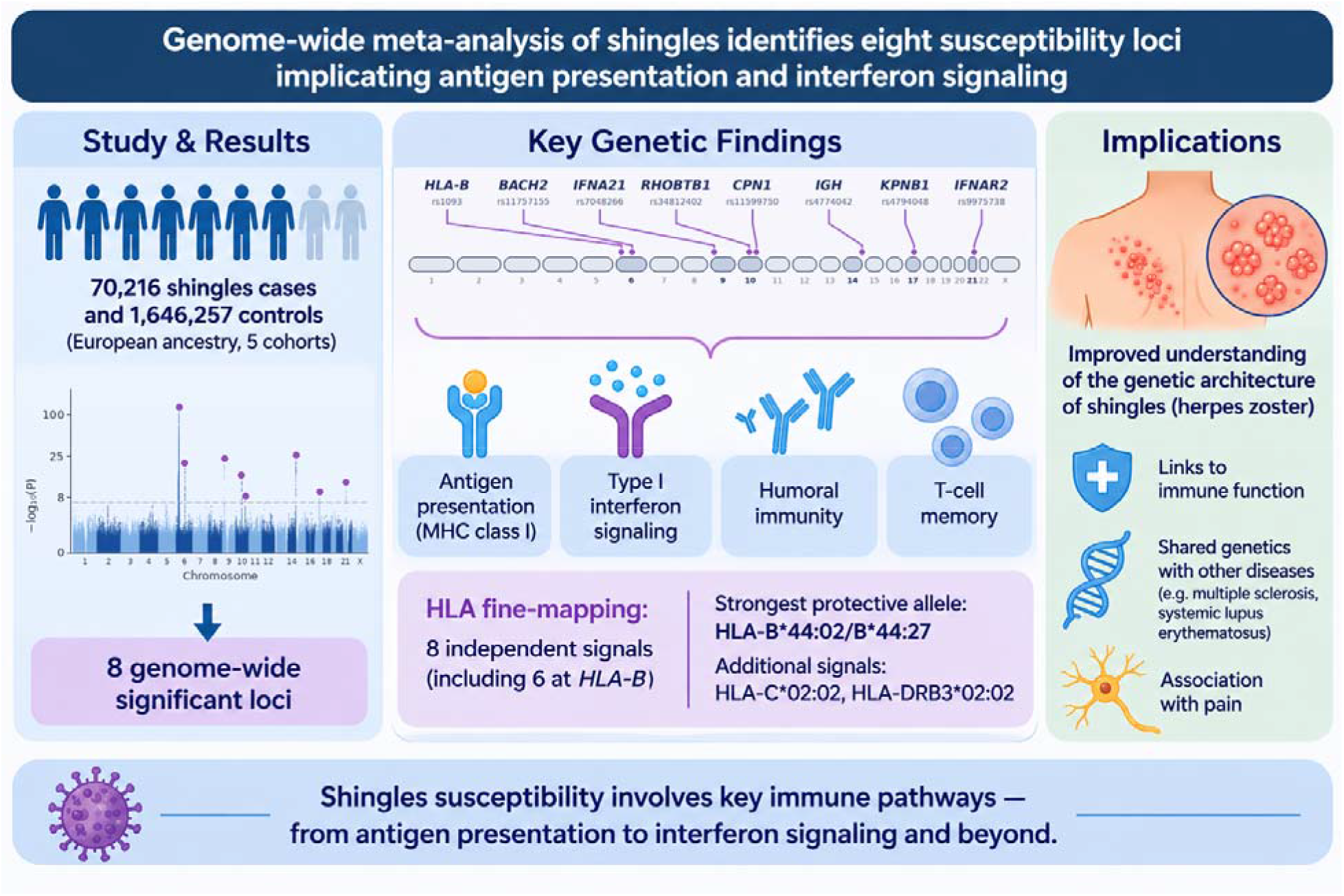

## Introduction

Shingles, also known as herpes zoster, is caused by reactivation of varicella zoster virus (VZV). Without vaccination, about one third of people develop shingles during their lifetime^1,2^. Vaccination reduces the risk by 50–90%, depending on the vaccine^3,4^, but shingles remains a major public health burden, partly because vaccine uptake among older adults is low^5^.

Primary VZV infection causes chickenpox, usually in childhood, after which the virus establishes lifelong latency in sensory ganglia. Upon reactivation, the virus travels along sensory nerves to the skin and causes a painful, one-sided blistering rash, often preceded by local burning pain, fever, headache and fatigue^6,7^. The rash usually resolves within weeks, but about 18% of adult patients develop postherpetic neuralgia, defined as pain persisting for more than 90 days^1,8^. Reactivation in the ophthalmic division of the trigeminal nerve can cause herpes zoster ophthalmicus, which can lead to vision loss^9^.

The main risk factors for shingles are older age and weakened immunity, including chemotherapy, autoimmune disease, HIV infection and long-term corticosteroid use, although in many patients no cause is found^10^. Latent VZV is controlled mainly by virus-specific T cells, and the decline of this response with age is thought to allow reactivation^11^.

Shingles has also been associated with later disease. Epidemiological studies report a nearly 30% higher risk of cardiovascular events, such as stroke and myocardial infarction, after shingles^12^, as well as associations with Parkinson’s disease and vascular dementia^13^. For all-cause dementia, the evidence is inconsistent^14,15^. Whether these associations reflect shared susceptibility or effects of the infection itself is unclear.

Family studies suggest that genetic factors contribute to shingles risk^16,17^, and genetic studies have identified associations in the HLA region^18–20^. The largest GWAS to date, with 16,711 cases, identified several signals near *HLA-A* and *HLA-B* and an association 500 kb upstream of *IFNA21*. Beyond these loci, the genetic architecture of shingles is largely unknown, including which immune pathways contribute to susceptibility and whether shingles shares genetic risk with the diseases associated with it.

Here, we performed a GWAS meta-analysis of shingles across five cohorts (FinnGen, the Estonian Biobank, UK Biobank, the Million Veteran Program (MVP) and All of Us), totaling 70,216 cases and 1,646,257 controls of European ancestry. We characterized the identified loci using HLA fine-mapping, statistical fine-mapping, colocalization with gene expression and gene set analysis. To study the relationships between shingles and other diseases, we performed a phenotype-wide association study (CodeWAS), genetic correlation analysis, an observational analysis of disease risk after shingles, and bidirectional two-sample Mendelian randomization.

## Results

### Large-scale GWAS meta-analysis identifies eight genome-wide significant loci for shingles susceptibility

To understand the role of genetics in susceptibility to shingles, we performed a meta-analysis of FinnGen (27,818 cases and 471,937 controls), the Estonian Biobank (13,128 cases and 192,920 controls), UK Biobank (14,173 cases and 384,864 controls), the Million Veteran Program (11,696 cases and 428,273 controls) and All of Us (3,401 cases and 168,263 controls) (Supplementary Figures 1–5). We only included participants of European ancestry. With 70,216 cases and 1,646,257 controls and 10,129,537 variants, we identified eight genome-wide significant loci associated with shingles (Figure 1, Table 1, Supplementary Tables S1 and S2, Supplementary Figure 6).

**Figure 1.**
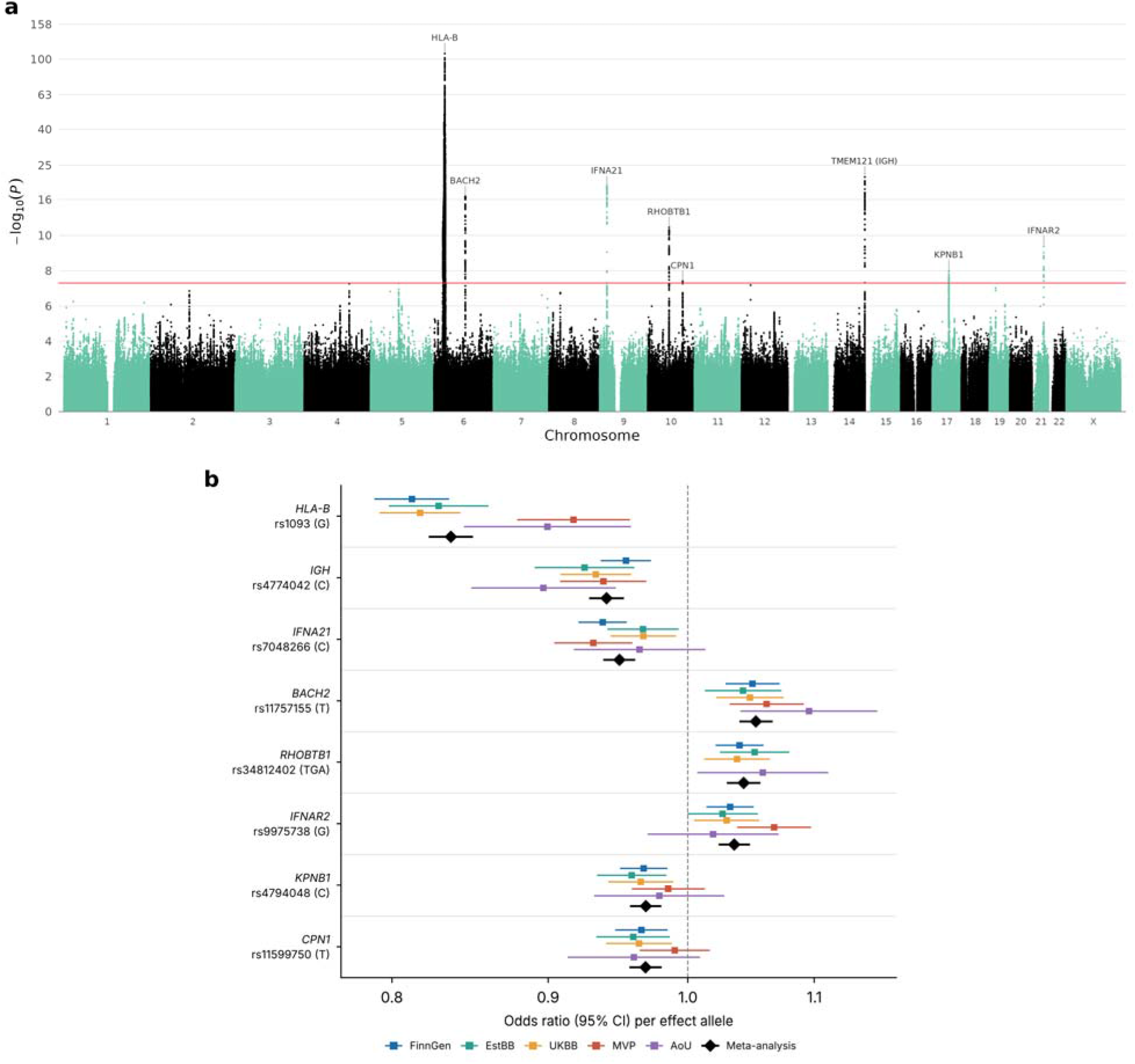
Shingles GWAS meta-analysis of five cohorts of European ancestry (70,216 cases and 1,646,257 controls). a, Manhattan plot. The red line indicates the genome-wide significance threshold (P = 5 × 10^−8^), loci are annotated with the nearest protein-coding gene (*TMEM121* lies in the *IGH* locus), and the y-axis is compressed (log scale) above −log_10_(P) = 10. b, Odds ratios (95% CI) of the lead variants in each cohort and in the meta-analysis, for the effect allele (Table 1). rs34812402 (*RHOBTB1*) was not available in MVP.

**Table 1.**
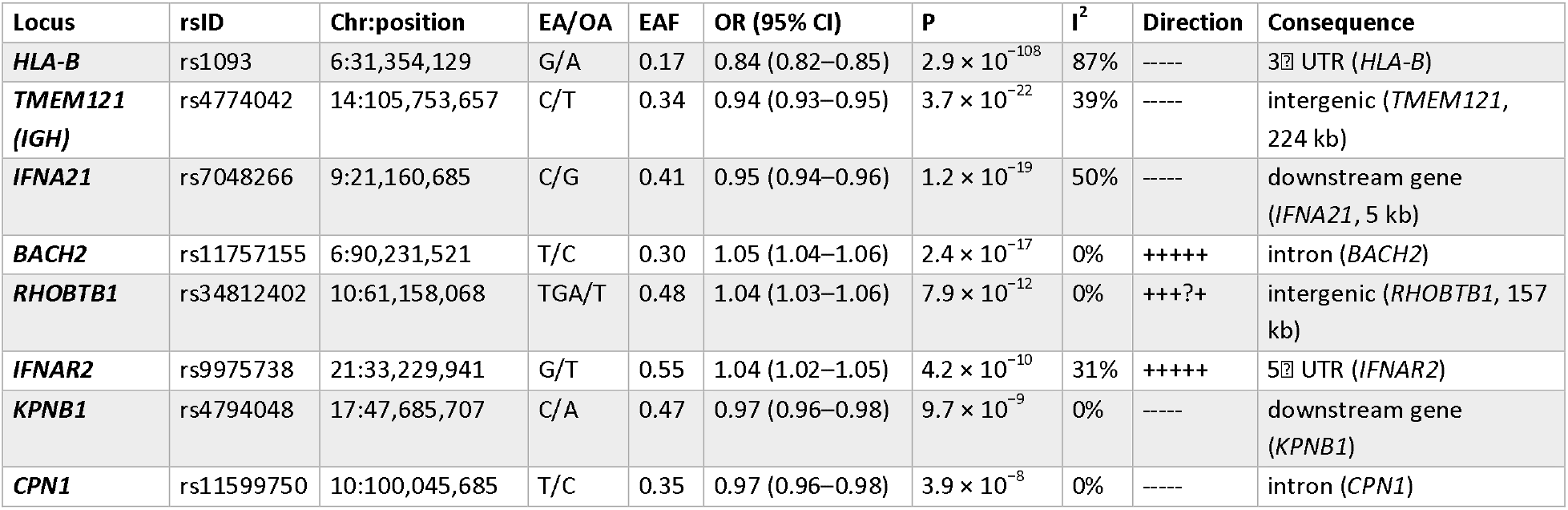
Lead variants of the shingles GWAS meta-analysis of five cohorts of European ancestry. Lead variants are COJO signals, except in the MHC, where the top variant is shown. Loci are named after the nearest protein-coding gene (*TMEM121* lies in the *IGH* locus). Positions are in GRCh38. The direction column shows the direction of effect in All of Us, EstBB, FinnGen, MVP and UKB (?, not available).

| Locus | rsID | Chr:position | EA/OA | EAF | OR (95% CI) | P | I<sup>2</sup> | Direction | Consequence |
|---|---|---|---|---|---|---|---|---|---|
| ***HLA-B*** | rs1093 | 6:31,354,129 | G/A | 0.17 | 0.84 (0.82–0.85) | 2.9 × 10<sup>−108</sup> | 87% | ----- | 3′ UTR (*HLA-B*) |
| ***TMEM121 (IGH)*** | rs4774042 | 14:105,753,657 | C/T | 0.34 | 0.94 (0.93–0.95) | 3.7 × 10<sup>−22</sup> | 39% | ----- | intergenic (*TMEM121*, 224 kb) |
| ***IFNA21*** | rs7048266 | 9:21,160,685 | C/G | 0.41 | 0.95 (0.94–0.96) | 1.2 × 10<sup>−19</sup> | 50% | ----- | downstream gene (*IFNA21*, 5 kb) |
| ***BACH2*** | rs11757155 | 6:90,231,521 | T/C | 0.30 | 1.05 (1.04–1.06) | 2.4 × 10<sup>−17</sup> | 0% | +++++ | intron (*BACH2*) |
| ***RHOBTB1*** | rs34812402 | 10:61,158,068 | TGA/T | 0.48 | 1.04 (1.03–1.06) | 7.9 × 10<sup>−12</sup> | 0% | +++?+ | intergenic (*RHOBTB1*, 157 kb) |
| ***IFNAR2*** | rs9975738 | 21:33,229,941 | G/T | 0.55 | 1.04 (1.02–1.05) | 4.2 × 10<sup>−10</sup> | 31% | +++++ | 5′ UTR (*IFNAR2*) |
| ***KPNB1*** | rs4794048 | 17:47,685,707 | C/A | 0.47 | 0.97 (0.96–0.98) | 9.7 × 10<sup>−9</sup> | 0% | ----- | downstream gene (*KPNB1*) |
| ***CPN1*** | rs11599750 | 10:100,045,685 | T/C | 0.35 | 0.97 (0.96–0.98) | 3.9 × 10<sup>−8</sup> | 0% | ----- | intron (*CPN1*) |

Conditional and joint analysis with COJO identified one independent signal at each of the seven loci outside the MHC (Supplementary Table S1). The lead variants were located in or near *HLA-B* (rs1093, OR = 0.84, P = 2.9 × 10^−108^), *TMEM121* in the immunoglobulin heavy chain (*IGH*) locus (rs4774042, OR = 0.94, P = 3.7 × 10^−22^), *IFNA21* (rs7048266, OR = 0.95, P = 1.2 × 10^−19^), *BACH2* (rs11757155, OR = 1.05, P = 2.4 × 10^−17^), *RHOBTB1* (rs34812402, OR = 1.04, P = 7.9 × 10^−12^), *IFNAR2* (rs9975738, OR = 1.04, P = 4.2 × 10^−10^), *KPNB1* (rs4794048, OR = 0.97, P = 9.7 × 10^−9^) and *CPN1* (rs11599750, OR = 0.97, P = 3.9 × 10^−8^). The direction of effect was consistent across all cohorts at every locus (Figure 1b). Although the SNP-based heritability of shingles was low (h² = 0.030 on the liability scale, SE = 0.003, assuming a lifetime prevalence of 30%, Supplementary Table S3), consistent with a large contribution of non-genetic factors such as age-related decline in VZV-specific immunity, the identified loci pointed to specific immune mechanisms, which we characterized with fine-mapping, colocalization and enrichment analyses.

Fine-mapping with SuSiE identified six 95% credible sets in six of the eight LD blocks outside the MHC (115 variants, Supplementary Table S4). The two credible sets at *RHOBTB1* were in adjacent LD blocks and their top variants were in LD (r^2^ = 0.73), consistent with the single COJO signal. The lead variant had the highest posterior inclusion probability (PIP) in its credible set at *IGH* (rs4774042, PIP = 0.72), *IFNAR2* (rs9975738, PIP = 0.34) and *RHOBTB1* (rs34812402, PIP = 0.32), whereas no variant had a PIP above 0.10 in the credible sets at *BACH2* and *IFNA21*. No credible set was identified at the *KPNB1* and *CPN1* loci.

The strongest signal was located in the *HLA-B* region, consistent with the well-established role of HLA class I variation in VZV immunity and CD8+ T cell-mediated control of viral reactivation^18,19,21^. Signals near *IFNA21* and *IFNAR2* highlight the importance of type I interferon signaling in shingles susceptibility. The *BACH2* signal suggests a role for T-cell regulation and memory, and the signal at the *IGH* locus points to antibody-mediated immunity.

Shingles risk increases with age (median age at first diagnosis in FinnGen, 66.9 years). Because the controls in our main analysis were population-based and included younger individuals, some of them may still develop shingles later in life, which could dilute the associations. Zoster vaccination similarly prevents shingles in individuals who might otherwise have become cases, and immunocompromise is a strong risk factor for shingles that could contribute to the genetic associations. To assess the influence of these factors, we repeated the GWAS in FinnGen restricting controls to individuals followed up to at least 70 years of age (27,818 cases and 187,102 controls), additionally excluding zoster-vaccinated controls (27,818 cases and 183,909 controls), and excluding cases and controls with severe immunocompromise (21,523 cases and 410,605 controls, Supplementary Table S5). The effects of the eight lead variants were consistent with the main FinnGen analysis in all three analyses. When their effect sizes were plotted against those from the main analysis, the regression slopes were 0.96–1.02, where a slope of 1 indicates identical effects (Supplementary Table S5). All lead variants remained associated (P ≤ 7.4 × 10^−3^) despite the smaller sample sizes. This suggests that population-based controls, as used in the meta-analysis, are suitable for studying the genetics of shingles susceptibility, and that the associations are not explained by control age, vaccination or immunocompromise.

### HLA fine-mapping identifies eight independent HLA allele signals

Our strongest association was in the MHC region, with the lead variant rs1093 located in the 3P UTR of *HLA-B*. Given the extensive LD in the region, we performed HLA fine-mapping in FinnGen using imputed HLA allele dosages (27,818 cases and 471,937 controls), where rs1093 had OR = 0.81 (P = 1.9 × 10^−49^). Of the 98 HLA alleles tested, 33 (17 protective and 16 risk alleles) were associated with shingles at the Bonferroni-corrected threshold (P < 5.1 × 10^−4^, Supplementary Table S6). Stepwise conditional analysis identified nine Bonferroni-significant independent alleles: B*44:02, C*02:02, B*35:01, B*18:01, B*44:27, B*15:01, DRB3*02:02, B*57:01 and B*27:05 (Supplementary Table S6).

B*44:02 and B*44:27 had similar protective effects (OR = 0.70 and 0.68 in the steps in which they were selected), so we tested whether they represent the same signal. In a model including both alleles and the other signals, their effects did not differ (P = 0.75), and the remaining B*44 alleles showed no association once B*44:02 and B*44:27 were accounted for (P = 0.51). We therefore combined the two alleles into one signal. In the joint model of the resulting eight signals, B*44:02/B*44:27 showed the strongest association (OR = 0.68 [0.65–0.72], P = 1.8 × 10^−61^), followed by C*02:02 (OR = 0.82, P = 1.5 × 10^−14^), B*35:01 (OR = 0.89, P = 1.0 × 10^−14^), B*18:01 (OR = 0.85, P = 4.6 × 10^−12^), B*15:01 (OR = 0.94, P = 3.2 × 10^−5^) and DRB3*02:02 (OR = 0.94, P = 2.2 × 10^−4^) (Figure 2, Supplementary Table S6). B*57:01 (OR = 1.16, P = 1.2 × 10^−4^) and B*27:05 (OR = 1.08, P = 4.3 × 10^−4^) were the only risk alleles. Six of the eight signals mapped to *HLA-B*. LD between the signals was low (r^2^ < 0.05), except between C*02:02 and B*27:05 (r^2^ = 0.38), which had opposite effects.

**Figure 2.**
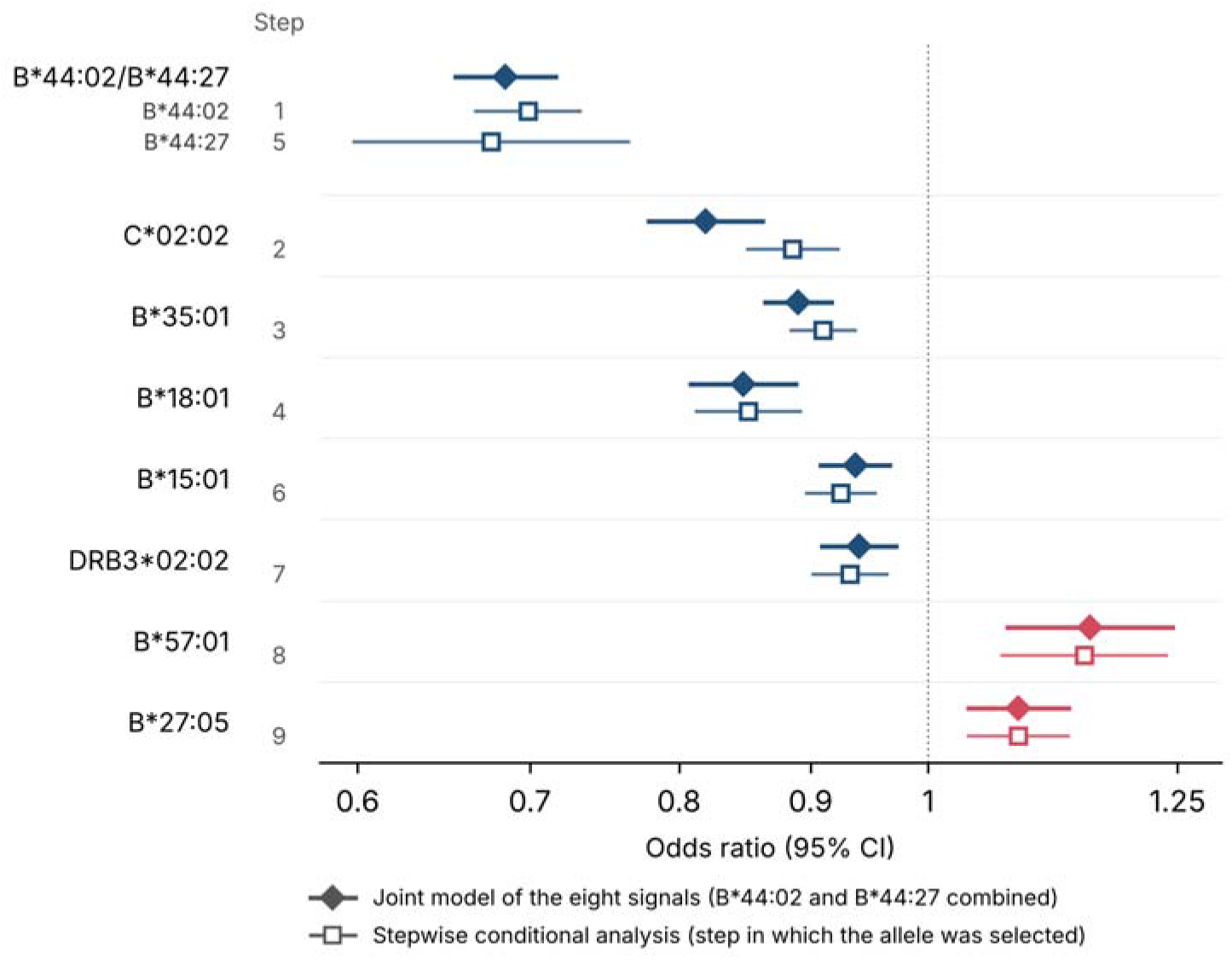
HLA alleles independently associated with shingles in FinnGen. Odds ratios and 95% confidence intervals from the joint model including all eight signals, in which B*44:02 and B*44:27 are combined into one signal, and from the stepwise conditional analysis (estimate in the step in which the allele was selected, conditioned on the alleles selected in earlier steps, with the step number on the left).

To verify our findings, we tested the HLA alleles in the Estonian Biobank without conditioning. Of the six signals available there (DRB3*02:02 and B*44:27 were not available), four replicated with the same direction of effect at P_FDR_ < 0.05: C*02:02, B*44:02, B*18:01 and B*57:01 (Supplementary Table S6).

### Colocalization links shingles loci to *IFNAR2*, *BACH2* and immunoglobulin gene expression

Gene-based analysis with MAGMA, excluding the extended MHC region, identified 14 genes at P < 2.5 × 10 (Bonferroni correction for 19,714 genes, Supplementary Table S7). Nine of these were at the GWAS loci: *KPNB1*, *NPEPPS*, *TBKBP1* and *TBX21* at the *KPNB1* locus, *IFNA21* and *IFNA7* at the *IFNA21* locus, and *BACH2*, *CPN1* and *IFNAR2* at their respective loci. The other five genes were in regions without a genome-wide significant variant, four on chromosome 5 (*MSH3*, *DHFR*, *MTRNR2L2*, *TERT*) and one on chromosome 15 (*CRTC3*).

To identify genes through which the loci outside the MHC region may act, we first looked up the lead variants and SuSiE credible-set variants in GTEx v10 (Supplementary Table S8). The variants at most loci were eQTLs for several genes, including *IFNAR2*, *IGHG1*, *IGHGP*, *DNMBP* and *TBKBP1* in whole blood, spleen or EBV-transformed lymphocytes. To test whether the GWAS and eQTL signals share a causal variant, we performed colocalization of the seven signals outside the MHC with 194 immune, skin and nerve eQTL datasets from the eQTL Catalogue (6,168 tests, Supplementary Table S9). In total, 27 gene–dataset pairs colocalized (PP.H4 ≥ 0.8), implicating ten genes at six loci (Figure 3, Supplementary Figure 7).

**Figure 3.**
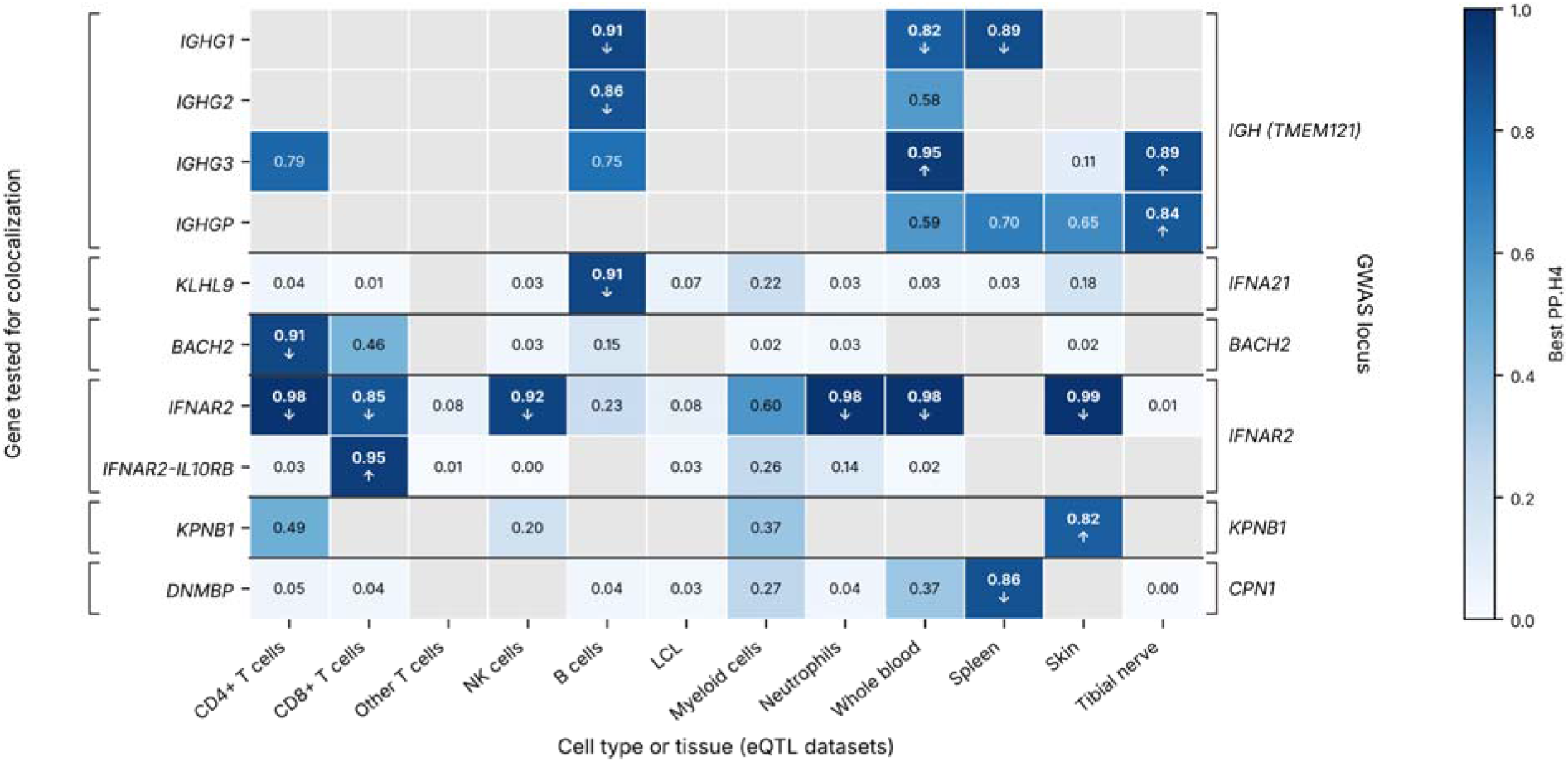
Colocalization of shingles GWAS signals with gene expression. Each cell shows the highest posterior probability of a shared causal variant (PP.H4) across the eQTL datasets in that cell type or tissue. Grey cells had no eQTL (P < 10) in the region. Bold values indicate colocalization (PP.H4 ≥ 0.8), and arrows show whether the shingles risk allele increases or decreases expression. Only genes that colocalized in at least one dataset are shown. LCL, EBV-transformed lymphocytes.

The strongest and most consistent evidence was at *IFNAR2*, where the shingles risk allele was associated with lower *IFNAR2* expression in CD4+ and CD8+ T cells, NK cells, neutrophils, whole blood and skin (PP.H4 = 0.85–0.99 in nine datasets). Other genes in the interferon receptor cluster, including *IFNAR1*, *IFNGR2* and *IL10RB*, had eQTLs driven by different causal variants (PP.H3 ≥ 0.8), suggesting that the signal acts through *IFNAR2*. At *BACH2*, the risk allele was associated with lower *BACH2* expression in CD4+ T cells and memory regulatory T cells (PP.H4 = 0.80–0.91 in five datasets). At the *IGH* locus, the signal colocalized with expression of the immunoglobulin heavy chain constant genes *IGHG1*, *IGHG2* and *IGHG3* and the pseudogene *IGHGP* in B cells, spleen, whole blood and tibial nerve (PP.H4 = 0.82–0.95), with opposite directions of effect for *IGHG1* and *IGHG3*.

Evidence at the remaining loci was weaker. Single datasets supported colocalization of the *IFNA21* signal with *KLHL9* expression in B cells (PP.H4 = 0.91), the *KPNB1* signal with *KPNB1* expression in skin (PP.H4 = 0.82), and the *CPN1* signal with *DNMBP* expression in spleen (PP.H4 = 0.86), but other datasets at the *IFNA21* and *CPN1* loci suggested distinct causal variants (PP.H3 ≥ 0.8 in 12 and 14 datasets, respectively). The *RHOBTB1* signal did not colocalize with any gene (maximum PP.H4 = 0.42).

Enhancer-to-gene (E2G) predictions supported the same genes at *IFNAR2* and *BACH2* (Supplementary Table S10). The *IFNAR2* lead variant rs9975738 lay in regulatory regions predicted to regulate *IFNAR2* (score 1.00), and LD proxies of the *BACH2* lead variant were linked to *BACH2* (score up to 1.00). At the other loci, E2G linked the signals to *IFNW1* (*IFNA21* locus), *TBKBP1*, *KPNB1*, *TBX21* and *NPEPPS* (*KPNB1* locus), *CPN1* and *DNMBP* (*CPN1* locus), *FAM30A* (*IGH* locus), and *LINC00845* and *TMEM26* (*RHOBTB1* locus), but we did not consider these predictions sufficient to assign target genes.

### Heritability enrichment and pathway analyses point to immune cells

To identify tissues and cell types relevant for shingles susceptibility, we used stratified LD score regression (sLDSC). Of the ten tissue groups, the immune group had the largest share of SNP heritability, with 23% of variants explaining 95% of the heritability (4.1-fold enrichment, P = 1.3 × 10^-5^). It was the only group whose annotation coefficient was positive beyond the baseline model (one-sided P = 0.014), although this did not pass Bonferroni correction for ten group (Supplementary Table S11). In the analysis of 205 gene expression and 489 chromatin annotations, no annotation reached P_FDR_ < 0.05, but the top annotations were immune: genes specifically expressed in NK cells, T cells, blood cells, EBV-transformed lymphocytes, whole blood and spleen (P = 5.9 × 10^-4^to 5.9 × 10^-3^), and H3K27ac, H3K4me1 and H3K4me3 marks in spleen, NK cells and T cells (P = 7.2 × 10^-4^to 2.0 × 10^-3^) (Supplementary Table S12).

MAGMA tissue expression analysis, excluding the extended MHC region, showed the same pattern. No tissue passed Bonferroni correction, and the strongest associations were with spleen (P = 2.8 × 10^−3^), EBV-transformed lymphocytes (P = 8.3 × 10^−3^) and whole blood (P = 0.034) among 54 GTEx tissues (Supplementary Table S7). Of 17,009 gene sets, only extrafollicular and follicular B cell activation (WikiPathways, 69 genes) passed Bonferroni correction (P = 3.5 × 10^−8^), and this association was driven by *BACH2*, *IFNAR2* and *TBX21*. When the MHC region was included, 190 genes passed Bonferroni correction (176 in the MHC), spleen became significant (P = 5.7 × 10⁻⁴), and the significant gene sets consisted mainly of HLA genes (Supplementary Table S7). Overall, sLDSC and MAGMA point to spleen and lymphocytes, with NK cells and T cells among the top sLDSC annotations, although most enrichments did not pass multiple-testing correction.

### Shingles is genetically correlated and co-occurs with pain, respiratory, infectious and immune-mediated diseases

Supporting a consistent case definition across the meta-analysis cohorts, shingles was strongly genetically correlated with the FinnGen zoster endpoint (rg = 0.93, P = 9.7 × 10⁻⁵⁶), which contains most of the cases in our FinnGen GWAS. Excluding the four endpoints that include shingles, shingles was genetically correlated with 290 FinnGen endpoints at P_FDR_ < 0.05 (Figure 4a, Supplementary Table S13). All of these genetic correlations were positive (rg = 0.12–0.59). The most significant genetic correlations were with pain conditions, including pain (rg = 0.39, P = 1.4 × 10⁻¹⁹), dorsalgia (rg = 0.36, P = 5.7 × 10⁻¹⁷) and joint pain (rg = 0.39, P = 1.1 × 10⁻¹⁶), and most of the other correlated endpoints were respiratory and digestive diseases (Figure 4a). Infectious diseases had some of the largest genetic correlations, including herpes simplex infection (rg = 0.59, P = 2.0 × 10⁻⁴), viral intestinal infections (rg = 0.50, P = 6.8 × 10⁻⁵) and COVID-19 (rg = 0.49, P = 1.5 × 10⁻³), although these estimates were less precise.

**Figure 4.**
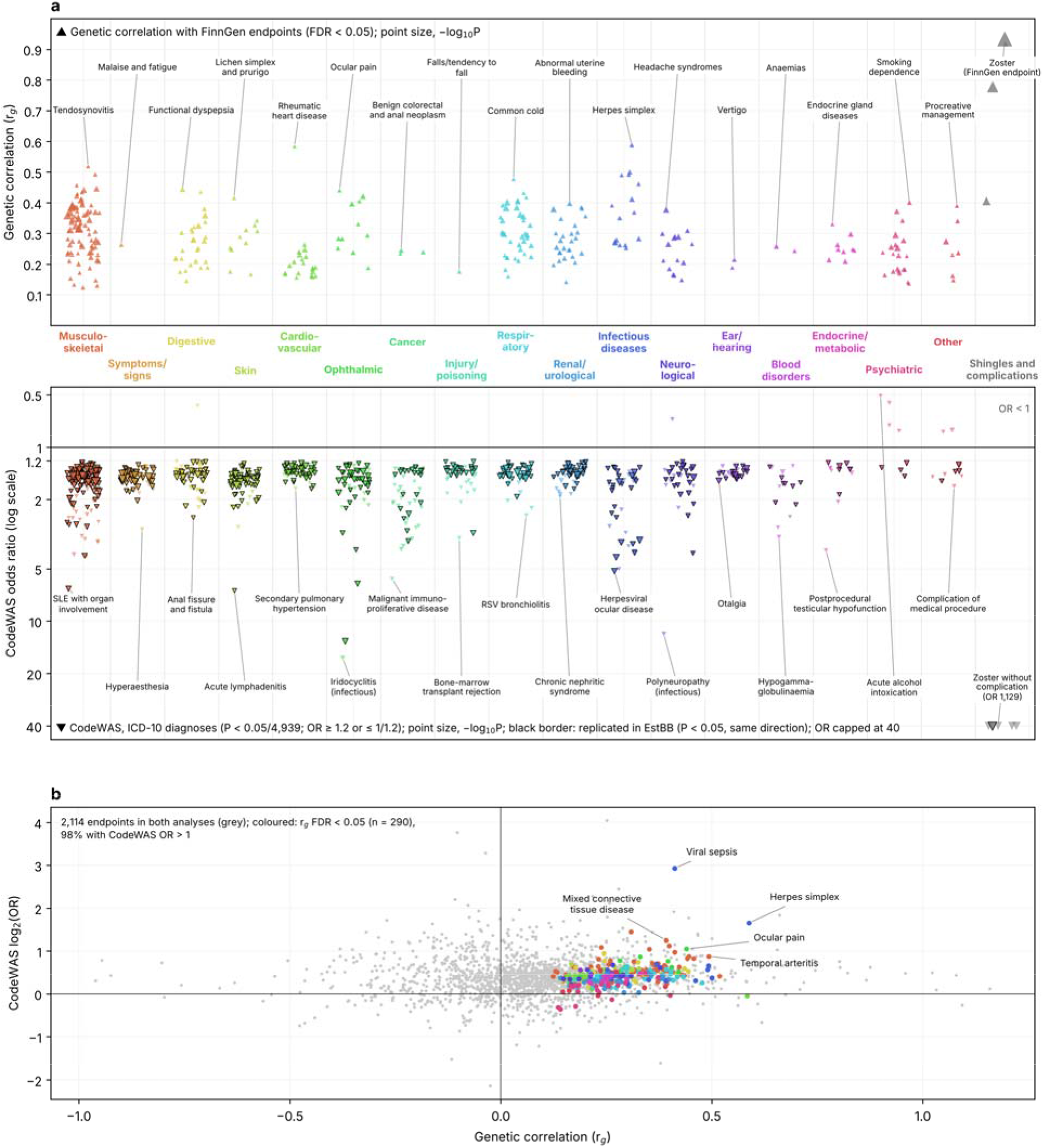
Genetic correlation and phenotype-wide associations of shingles with other diseases. a, Upper half: genetic correlation (rg, LD score regression) between the shingles meta-analysis and FinnGen R13 endpoints with P_FDR_ < 0.05. Point size is proportional to −log10 P. The shingles column shows the four FinnGen endpoints that include zoster (two zoster endpoints, viral skin and mucous membrane infections, and all infectious and parasitic diseases). The other columns show the 290 endpoints that do not. Lower half: diagnoses associated with shingles in the FinnGen CodeWAS (27,818 cases and 201,350 matched controls). ICD-10 codes with P < 1.0 × 10⁻⁵ and OR ≥ 1.2 or ≤ 1/1.2 are shown. Odds ratios are on a log scale and capped at 40. Black borders mark associations replicated in the Estonian Biobank (P < 0.05, same direction). Endpoints are grouped by FinnGen endpoint category and codes by ICD-10 chapter, with zoster codes and complications as a separate group (Supplementary Tables S13 and S14), and the trait with the largest effect (rg or OR) in each group is named. b, Genetic correlation against CodeWAS odds ratio for the 2,114 FinnGen endpoints tested in both analyses, excluding endpoints that include shingles. Colored points, rg P_FDR_ < 0.05 (n = 290). The five endpoints furthest to the upper right are named.

In the CodeWAS of 27,818 cases and 201,350 age-and sex-matched controls in FinnGen, 798 of 4,939 ICD-10 diagnosis codes were associated with shingles (P < 1.0 × 10⁻⁵ and OR ≥ 1.2 or ≤ 1/1.2; Figure 4a, Supplementary Table S14). Supporting the case definition, zoster codes were strongly enriched in cases (OR = 1,129 for zoster without complication, 336 for zoster ocular disease, 276 for zoster with other complications and 53 for postzoster neuralgia). Of the other 793 associated codes, 785 were more common in cases. The strongest associations were with pain diagnoses (OR = 1.58, P = 1.3 × 10⁻²⁷² for pain in limb and OR = 1.47, P = 2.2 × 10⁻¹⁷⁷ for low back pain) and common skin and respiratory infections. The largest odds ratios were for infection-related iridocyclitis (OR = 16.3) and keratitis (OR = 13.1), which likely include zoster complications, other herpesvirus infections (OR = 5.2 for herpesviral ocular disease and 4.3 for eczema herpeticum), immunodeficiency and transplantation (OR = 3.3 for hypogammaglobulinemia and 3.1 for failure or rejection of transplanted organs), chronic lymphocytic leukemia (OR = 2.4), SLE with organ involvement (OR = 6.5) and multiple sclerosis (OR = 1.65). The eight codes less common in cases included Alzheimer’s disease (OR = 0.69), schizophrenia (OR = 0.56) and alcohol-related disorders. Of the 718 associated codes tested in the Estonian Biobank, 540 (75%) replicated with the same direction of effect (P < 0.05) and 284 passed Bonferroni correction (P < 0.05/5,508).

The clinical and genetic associations agreed. Of the 290 FinnGen endpoints genetically correlated with shingles, 283 (98%) were also more common in shingles cases in the CodeWAS (Figure 4b).

### Observational associations and Mendelian randomization

Shingles cases were more often diagnosed than matched controls with herpes simplex (OR = 3.24 [3.07–3.42]), SLE (OR = 2.54 [2.17–2.97]), multiple sclerosis (OR = 1.75 [1.53–2.01]), giant cell arteritis (OR = 1.69 [1.46–1.95]) and Bell’s palsy (OR = 1.52 [1.38–1.67]) (Figure 5a, Supplementary Table S15). These associations were present both before and after shingles, and new-onset herpes simplex (OR = 25.1 [20.0–31.6]) and Bell’s palsy (OR = 15.2 [10.3–22.3]) were particularly common within 90 days after shingles. Myocardial infarction (OR = 1.17 [1.12– 1.23]) and stroke (OR = 1.08 [1.03–1.13]) were also more common in cases, but so was the negative control outcome, foot or ankle fracture (OR = 1.15 [1.10–1.19]), suggesting residual confounding of this size. Within 90 days after shingles, new-onset stroke (OR = 1.59 [1.20– 2.10]) and myocardial infarction (OR = 1.52 [1.09–2.12]) were more common, whereas fracture was not (OR = 1.17 [0.82–1.68]). Dementia (OR = 0.84 [0.80–0.88]) and Alzheimer’s disease (OR = 0.81 [0.77–0.86]) were less common in shingles cases, and Parkinson’s disease and Guillain– Barré syndrome were not associated.

**Figure 5.**
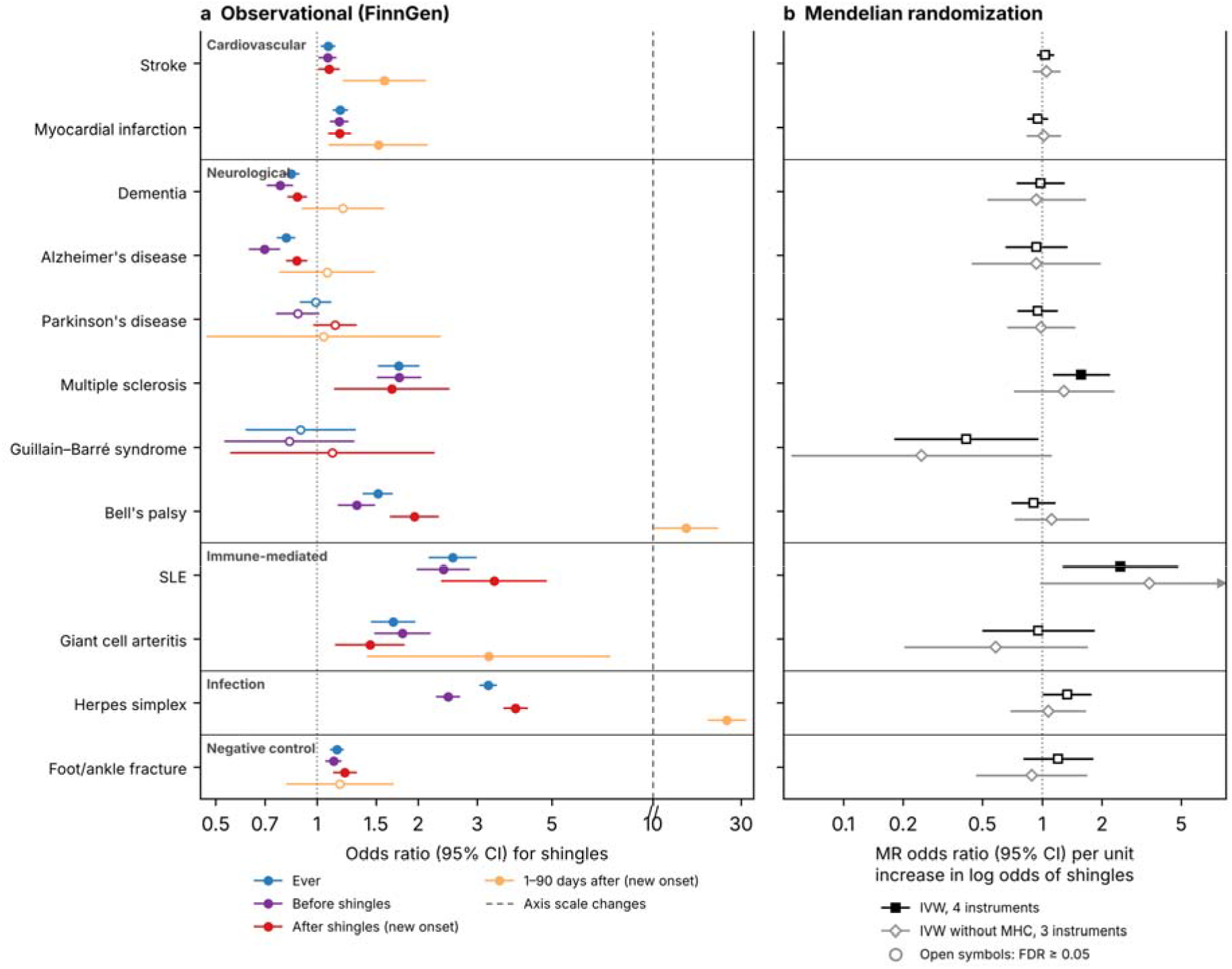
Observational and genetic associations of shingles with the Mendelian randomization outcomes. a, Odds ratios (95% CI) from conditional logistic regression in FinnGen comparing 27,724 shingles cases with 244,315 matched controls, for the outcome recorded ever, before shingles, after shingles (new onset) and 1–90 days after shingles (new onset). The x-axis i compressed above 10 (dashed line). Filled symbols, P_FDR_ < 0.05 within the time window. b, Inverse-variance weighted MR estimates of the effect of genetic liability to shingles on each outcome, with four genome-wide significant instruments (one in the MHC) and without the MHC instrument (three instruments). Open symbols, P_FDR_ ≥ 0.05.

In two-sample MR with four genome-wide significant instruments, genetic liability to shingles was associated with multiple sclerosis (OR = 1.57 [1.15–2.16], P_FDR_ = 0.041) and SLE (OR = 2.47 [1.28–4.76], P_FDR_ = 0.041) (Figure 5b, Supplementary Table S16). Both associations depended on the MHC instrument. In a sensitivity analysis without it, neither was significant (OR = 1.29 [0.73–2.28], P = 0.38 for multiple sclerosis and OR = 3.46 [0.99–12.1], P = 0.052 for SLE). Herpes simplex (OR = 1.33 [1.02–1.75], P_FDR_ = 0.11) and Guillain–Barré syndrome (OR = 0.42 [0.18– 0.95], P_FDR_ = 0.11) did not pass FDR correction, and we found no evidence of a causal effect of genetic liability to shingles on stroke, myocardial infarction, dementia, Alzheimer’s disease, Parkinson’s disease, Bell’s palsy, giant cell arteritis or the negative control. The MR-Egger intercept did not indicate directional pleiotropy for any outcome (all P > 0.1).

In the reverse direction, we tested genetic liability to 334 FinnGen disease endpoints a exposures (Supplementary Table S17). Four were associated with shingles at P_FDR_ < 0.05: pain (OR = 1.22 [1.12–1.33], P_FDR_ = 7.4 × 10⁻⁴), seborrhoeic keratosis (OR = 1.08 [1.05–1.12], P_FDR_ = 4.9 × 10⁻⁴), and inversely thyroid cancer (OR = 0.96 [0.95–0.98], P_FDR_ = 0.013) and the broader endpoint of endocrine gland cancers (OR = 0.96 [0.94–0.98], P_FDR_ = 0.013). For all four, the weighted median estimates had the same direction and similar size (all P ≤ 1.6 × 10⁻³), MR-PRESSO detected no outliers, and no single instrument drove the associations in leave-one-out analyses. We found no evidence that genetic liability to multiple sclerosis (OR = 1.01 [0.97– 1.05], P_FDR_ = 0.88) or SLE (OR = 1.01 [0.98–1.03], P_FDR_ = 0.92) affects the risk of shingles. Genetic liability to dementia was nominally associated with a lower risk of shingles, but this did not pas FDR correction (OR = 0.98 [0.97–1.00], P_FDR_ = 0.28).

## Discussion

In this meta-analysis of 70,216 shingles cases and 1,646,257 controls, we identified eight genome-wide significant loci. Together with fine-mapping and colocalization, these loci implicate four parts of immunity in shingles susceptibility: antigen presentation by HLA class I molecules, type I interferon signaling, T-cell regulation and memory, and antibody-mediated immunity.

The strongest association was in the MHC, where HLA fine-mapping identified eight independent signals: six at *HLA-B*, one at *HLA-C* (C*02:02) and one in class II (DRB3*02:02). Most were protective, led by HLA-B*44:02/B*44:27, whereas B*57:01 and B*27:05 increased risk. Four of the signals replicated in the Estonian Biobank. HLA associations with shingles have been reported before^18–20^ and are consistent with our findings. Of our eight signals, the protective B*44:02 and the risk allele B*57:01 were also reported by Tian et al.^19^, and DRB3*02:02 was protective in UK Biobank^20^.

HLA class I molecules present viral peptides to CD8⁺ T cells, a pathway that VZV actively suppresses^22^, and the allele-specific effects at *HLA-B* likely reflect differences in the presentation of VZV peptides. However, VZV-specific CD8⁺ T cells are rare in latently infected trigeminal ganglia^23^, suggesting a limited role in maintaining latency, whereas CD8⁺ and CD4⁺ T cells infiltrate ganglia during reactivation^21^. CD8⁺ T-cell responses through HLA class I may therefore matter mainly after the virus reactivates, rather than in keeping it latent. The class II signal at DRB3*02:02 points to CD4⁺ T cells. In a large vaccine trial, stronger VZV-specific cell-mediated immunity at shingles onset was associated with milder disease and less postherpetic neuralgia^24^.

Two loci implicate type I interferon signaling. The *IFNA21* signal replicates the association reported by Tian et al.^19^, whose lead variant rs7047299 is in strong LD with our lead variant rs7048266 (r^2^ = 0.98 in 1000 Genomes EUR), with a consistent direction of effect. The mechanism at this locus remains unclear. The signal lies in the type I interferon gene cluster, enhancer–gene predictions linked it to *IFNW1*, and colocalization suggested a shared variant with *KLHL9* expression in only one B cell dataset. In contrast, the *IFNAR2* locus has a clear candidate gene. *IFNAR2* encodes the high-affinity subunit of the type I interferon receptor, through which type I interferons restrict VZV replication^25^. The shingles risk allele was associated with lower *IFNAR2* expression in nine datasets, including T cells, NK cells, neutrophils, whole blood and skin, whereas eQTLs of the neighboring receptor genes *IFNAR1*, *IFNGR2* and *IL10RB* were driven by different variants. Complete *IFNAR2* deficiency causes life-threatening disease after live-attenuated viral vaccination^26^, and herpes simplex virus 1 (HSV-1), a close relative of VZV, blocks type I interferon signaling^27^. Our findings suggest that common variation in interferon receptor expression also affects control of latent VZV. Together, the *IFNA21* and *IFNAR2* loci implicate both interferon production and receptor signaling.

At *BACH2*, the risk allele was associated with lower *BACH2* expression in CD4⁺ T cells and memory regulatory T cells. *BACH2* is a transcription factor that helps T cells become long-lived memory cells rather than short-lived effector cells^28^, and it also stabilizes regulatory T cells^29^. In humans, loss of one functional copy of *BACH2* causes an immunodeficiency with impaired lymphocyte maturation and recurrent infections^30^. Lower *BACH2* expression could therefore weaken VZV-specific memory T cells, which become less functional with age^31^. Common variants at *BACH2* have also been associated with several autoimmune diseases^30^, which is of interest given the epidemiological and genetic links between shingles and SLE observed here.

At the *TMEM121* (*IGH*) locus, the lead variant lies within the immunoglobulin heavy chain constant region, and the signal colocalized with the expression of immunoglobulin G constant genes (*IGHG1*, *IGHG2*, *IGHG3* and the pseudogene *IGHGP*) rather than *TMEM121*, pointing to humoral immunity. The lead variant was in strong LD (r^2^ = 0.89) with the lead variant for serum IgG1 levels in a recent GWAS (rs74093865^32^). This variant tags G1m(f), the most common IgG1 allotype. The shingles risk allele tagged the allele associated with lower IgG1 levels, consistent with the lower *IGHG1* expression observed here. In the same study, this haplotype was associated with higher IgG3, matching the opposite directions of effect we observed for *IGHG1* and *IGHG3*, so the locus may shift the IgG subclass balance rather than total IgG. IgG1 is the dominant VZV-specific IgG subclass during reactivation^33^, and low VZV-specific IgG1 together with absent IgG3 has been proposed to explain shingles susceptibility in immunodeficient patients^34^.

To our knowledge, the *KPNB1*, *CPN1* and *RHOBTB1* loci have not previously been associated with shingles, and their causal genes remain unresolved. The *KPNB1* locus contains three genes with roles in antiviral immunity. *KPNB1* encodes importin-β1, which carries the interferon-activated transcription factor STAT1 into the nucleus^35^ and is used by herpes simplex virus 1 (HSV-1) to deliver its genome^36^. *TBKBP1* encodes an adaptor of the kinase TBK1, which induces type I interferons^37^, and *TBX21* encodes T-bet, which is required for Th1 and NK cell responses^38^. Colocalization supported only *KPNB1*, in a single skin dataset. At the *CPN1* locus, the lead variant is intronic to *CPN1*, which encodes a complement-inactivating enzyme^39^, but the signal colocalized with *DNMBP* expression in spleen. The *RHOBTB1* signal did not colocalize with any gene.

At the pathway level, the analyses consistently pointed to immune cells. In sLDSC, immune annotations covered 23% of variants but explained 95% of the SNP heritability (4.1-fold enrichment), with NK cells and T cells as the top cell types. In MAGMA, spleen and lymphocyte expression ranked highest, and spleen became significant when the MHC was included. Among gene sets, B cell activation was significant, driven by *BACH2*, *IFNAR2* and *TBX21*.

Beyond the genetic loci, shingles was linked to a wide range of other diseases. In FinnGen, almost 800 diagnosis codes were more common in shingles cases than in matched controls, three quarters of those tested replicated in the Estonian Biobank, and shingles was positively genetically correlated with 290 FinnGen endpoints. The clinical and genetic associations largely agreed, as nearly all genetically correlated endpoints were also more common in shingles cases. Part of this broad pattern likely reflects factors that affect many diagnoses at once, such as healthcare use, general health and immune function. The negative control outcome, foot or ankle fracture, was 15% more common in shingles cases, which indicates the size of such confounding in the observational analyses.

Pain conditions had the strongest genetic correlations with shingles, and in reverse MR, with the FinnGen pain endpoint as the exposure and shingles as the outcome, genetic liability to pain was associated with a higher risk of shingles. Shingles often begins with pain in the affected dermatome before the rash appears, and pain is also its most common complication. The shared genetic basis could reflect differences in pain sensitivity or shared biology in sensory neurons, where VZV establishes latency. The associations with genetic liability to seborrhoeic keratosis and, inversely, with thyroid cancer have no clear explanation and need replication.

Among the twelve outcomes selected for Mendelian randomization, the strongest observational associations were with immune-mediated diseases and herpes simplex. In this analysis, which compared shingles cases with matched controls before and after the shingles diagnosis, SLE and multiple sclerosis were more common in cases in both periods. In forward MR, with shingles as the exposure, genetic liability to shingles was associated with a higher risk of both diseases, but these associations were driven by the MHC instrument and were not significant without it. Because HLA alleles strongly affect autoimmune diseases, this likely reflects shared HLA effects rather than a causal effect of shingles. The observational associations, already present before shingles, are more consistent with these diseases or their treatment increasing the risk of shingles, as reported for SLE^40,41^ and for several multiple sclerosis therapies^42^. Herpes simplex had the highest genetic correlation with shingles and a strong observational association, consistent with shared host control of latent alphaherpesviruses. Misclassification between zosteriform herpes simplex and shingles may also contribute, particularly shortly after shingles.

Shingles has been associated with an increased risk of stroke and myocardial infarction, particularly shortly after the infection^12^. We observed the same pattern. New-onset stroke and myocardial infarction were more common within 90 days after shingles, whereas over longer periods the associations were of similar size to that of the negative control. MR did not support a causal effect of genetic liability to shingles on stroke or myocardial infarction. Together, these results are consistent with a transient effect of the acute infection, which MR of lifelong genetic liability cannot capture, rather than a long-term effect of susceptibility to shingles. Bell’s palsy was very common shortly after shingles, which likely reflects facial palsy caused by VZV itself (Ramsay Hunt syndrome) being recorded as Bell’s palsy. Dementia and Alzheimer’s disease were less common in shingles cases, both before and after shingles, and MR did not support a causal effect of shingles on dementia. This inverse association may reflect under-diagnosis of shingles in people with dementia, who may be less able to report symptoms, and illustrates why observational studies of shingles and dementia have given inconsistent results^14,15^.

Our study has limitations. First, all cohorts were of European ancestry, and the findings need to be tested in other populations. Second, shingles was defined from diagnosis codes in health registries and electronic health records. Not all patients seek care, data sources and coding practices differed between cohorts, and misclassification would bias the associations towards the null. Registry-recorded cases made up only 2–6% of each cohort, much less than the assumed lifetime prevalence of 30%, likely because many participants have not yet reached the ages of highest risk and health records cover only part of each participant’s life. The liability-scale heritability estimates (Supplementary Table S3) depend on prevalence assumptions and need to be interpreted with caution. Third, conditional analysis and SuSiE fine-mapping used 1000 Genomes European samples as the LD reference, which does not include Finns, although FinnGen contributed 40% of the cases, and LD differences may affect these results. Fourth, colocalization assumed a single causal variant per signal, so genes at loci with several causal variants may have been missed. Fifth, sequential HLA fine-mapping was performed in FinnGen only, at two-field resolution. In the Estonian Biobank, HLA alleles were imputed with a different method and tested one at a time, without sequential conditioning. Sixth, the CodeWAS compared lifetime diagnoses with matching only on age and sex, so its associations describe co-occurrence and are subject to confounding. Finally, MR had only four instruments for shingles, three outside the MHC, which limits power and the ability to separate causal effects from pleiotropy.

In conclusion, we identified eight loci for shingles, including six not previously reported, and our results implicate HLA class I antigen presentation, type I interferon signaling, T-cell memory and antibody-mediated immunity in the control of VZV reactivation. Shingles shares clinical and genetic links with pain, infectious and immune-mediated diseases, but MR did not support causal effects of genetic liability to shingles on cardiovascular, neurodegenerative or autoimmune diseases beyond shared HLA effects. These findings improve understanding of host control of VZV latency and provide candidate pathways for further functional studies.

## Materials and methods

### Cohorts

The FinnGen study is a large-scale genomics initiative that has analyzed over 500,000 Finnish biobank samples and correlated genetic variation with health data to understand disease mechanisms and predispositions^43^. The project is a collaboration between research organizations and biobanks within Finland and international industry partners. Shingles cases were identified from longitudinal health registry data using ICD-10 code B02 and ICD-8 and ICD-9 code 053 from inpatient, outpatient and primary care registries. The analysis included 27,818 cases and 471,937 controls.

The Estonian Biobank (EstBB) is a large population-based biobank comprising 212,955 participants^44^. Electronic health record data, including ICD-10 diagnostic codes, are obtained through regular linkage with the National Health Insurance Fund and other relevant databases, with the majority of records available from 2004 onwards^45^. Shingles cases were identified using ICD10 code B02, using data freeze 2026v02.

The UK Biobank (UKB) is a large prospective open-access cohort study that recruited over 500,000 individuals aged 40–69 years between 2006 and 2010. At enrollment, participants provided blood and urine samples for genetic and biochemical analysis, alongside detailed health and lifestyle measures. Hospital inpatient (HES; N ∼ 470,000) and primary care (GP; N ∼ 231,000) records were subsequently linked to provide longitudinal data on disease diagnoses, operations, medications, and deaths. In this study, we used publicly available summary statistics for Zoster [herpes zoster] from the European subset of the pan-UKB study, provided through FinnGen’s FG-UKB-MVP meta-analysis resource (https://mvp-ukbb.finngen.fi).

The Million Veteran Program (MVP) is a large longitudinal cohort study examining how genes, lifestyle, military experiences, and exposures influence health among diverse U.S. Veterans. It combines genetic data with electronic health records from 635,969 participants across four ancestral groups. In this study, we used summary statistics obtained through FinnGen’s meta-analysis resource for Zoster [herpes zoster] from European ancestry (https://mvp-ukbb.finngen.fi).

The All of Us Research Program is a large prospective cohort study launched by the National Institutes of Health (NIH) in 2018, aiming to enroll one million or more diverse participants across the United States. The program collects a broad range of data including genetic information, electronic health records, physical measurements, and lifestyle and environmental factors, with a particular emphasis on diversity and inclusion of historically underrepresented populations in biomedical research. In this study, we used summary statistics obtained through FinnGen’s FG-AoU meta-analysis resource (https://metaresults-aou.finngen.fi) for Zoster [herpes zoster] from European ancestry.

### Ethics statements

Study subjects in FinnGen provided informed consent for biobank research, based on the Finnish Biobank Act. Alternatively, separate research cohorts, collected prior the Finnish Biobank Act came into effect (in September 2013) and start of FinnGen (August 2017), were collected based on study-specific consents and later transferred to the Finnish biobanks after approval by Fimea (Finnish Medicines Agency), the National Supervisory Authority for Welfare and Health. Recruitment protocols followed the biobank protocols approved by Fimea. The Coordinating Ethics Committee of the Hospital District of Helsinki and Uusimaa (HUS) statement number for the FinnGen study is Nr HUS/990/2017.

The FinnGen study is approved by Finnish Institute for Health and Welfare (permit numbers: THL/2031/6.02.00/2017, THL/1101/5.05.00/2017, THL/341/6.02.00/2018, THL/2222/6.02.00/2018, THL/283/6.02.00/2019, THL/1721/5.05.00/2019 and THL/1524/5.05.00/2020), Digital and population data service agency (permit numbers: VRK43431/2017-3, VRK/6909/2018-3, VRK/4415/2019-3), the Social Insurance Institution (permit numbers: KELA 58/522/2017, KELA 131/522/2018, KELA 70/522/2019, KELA 98/522/2019, KELA 134/522/2019, KELA 138/522/2019, KELA 2/522/2020, KELA 16/522/2020), Findata permit numbers THL/2364/14.02/2020, THL/4055/14.06.00/2020, THL/3433/14.06.00/2020, THL/4432/14.06/2020, THL/5189/14.06/2020, THL/5894/14.06.00/2020, THL/6619/14.06.00/2020, THL/209/14.06.00/2021, THL/688/14.06.00/2021, THL/1284/14.06.00/2021, THL/1965/14.06.00/2021, THL/5546/14.02.00/2020, THL/2658/14.06.00/2021, THL/4235/14.06.00/2021, Statistics Finland (permit numbers: TK-53-1041-17 and TK/143/07.03.00/2020 (earlier TK-53-90-20) TK/1735/07.03.00/2021, TK/3112/07.03.00/2021) and Finnish Registry for Kidney Diseases permission/extract from the meeting minutes on 4th July 2019.

The Biobank Access Decisions for FinnGen samples and data utilized in FinnGen Data Freeze 14 include: THL Biobank BB2017_55, BB2017_111, BB2018_19, BB_2018_34, BB_2018_67, BB2018_71, BB2019_7, BB2019_8, BB2019_26, BB2020_1, BB2021_65, Finnish Red Cross Blood Service Biobank 7.12.2017, Helsinki Biobank HUS/359/2017, HUS/248/2020, HUS/430/2021_28, _29, HUS/150/2022 _12, _13, _14, _15, _16, _17, _18, _23, _58, _59, HUS/128/2023 _18, Auria Biobank AB17-5154 and amendment #1 (August 17 2020) and amendments BB_2021-0140, BB_2021-0156 (August 26 2021, Feb 2 2022), BB_2021-0169, BB_2021-0179, BB_2021-0161, AB20-5926 and amendment #1 (April 23 2020) and it’s modifications (Sep 22 2021), BB_2022-0262, BB_2022-0256, Biobank Borealis of Northern Finland_2017_1013, 2021_5010, 2021_5010 Amendment, 2021_5018, 2021_5018 Amendment, 2021_5015, 2021_5015 Amendment, 2021_5015 Amendment_2, 2021_5023, 2021_5023 Amendment, 2021_5023 Amendment_2, 2021_5017, 2021_5017 Amendment, 2022_6001, 2022_6001 Amendment, 2022_6006 Amendment, 2022_6006 Amendment, 2022_6006 Amendment_2, BB22-0067, 2022_0262, 2022_0262 Amendment, Biobank of Eastern Finland 1186/2018 and amendment 22_/2020, 53_/2021, 13_/2022, 14_/2022, 15_/2022, 27_/2022, 28_/2022, 29_/2022, 33_/2022, 35_/2022, 36_/2022, 37_/2022, 39_/2022, 7_/2023, 32_/2023, 33_/2023, 34_/2023, 35_/2023, 36_/2023, 37_/2023, 38_/2023, 39_/2023, 40_/2023, 41_/2023, Finnish Clinical Biobank Tampere MH0004 and amendments (21.02.2020 & 06.10.2020), BB2021-0140 8_/2021, 9_/2021, _9/2022, _10/2022, _12/2022, 13_/2022, _20/2022, _21/2022, _22/2022, _23/2022, 28_/2022, 29_/2022, 30_/2022, 31_/2022, 32_/2022, 38_/2022, 40_/2022, 42_/2022, 1_/2023, Central Finland Biobank 1-2017, BB_2021-0161, BB_2021-0169, BB_2021-0179, BB_2021-0170, BB_2022-0256, BB_2022-0262, BB22-0067, Decision allowing to continue data processing until 31st Aug 2024 for projects: BB_2021-0179, BB22-0067,BB_2022-0262, BB_2021-0170, BB_2021-0164, BB_2021-0161, and BB_2021-0169, and Terveystalo Biobank STB 2018001 and amendment 25th Aug 2020, Finnish Hematological Registry and Clinical Biobank decision 18th June 2021, Arctic biobank P0844: ARC_2021_1001.

The activities of the EstBB are regulated by the Human Genes Research Act, which was adopted in 2000 specifically for the operations of EstBB. Individual level data analysis in EstBB was carried out under ethical approval 1.1-12/624 from the Estonian Committee on Bioethics and Human Research (Estonian Ministry of Social Affairs), using data according to release application 6-7/GI/10314 from the Estonian Biobank.

### Genotyping and quality control

FinnGen R14 contains genetic data for 519,972 individuals. The samples were genotyped using Illumina (Illumina) and Affymetrix arrays (Thermo Fisher Scientific). The array consisted of 735,145 probes looking for 655,973 variants consisting of core backbone variants for imputation, rare coding variants enriched in the Finnish population, variants for KIR and HLA haplotypes, disease-specific markers and pharmacogenomic markers. Genotyping data produced with previous chip platforms and reference genome builds were lifted over to build v.38 (GRCh38/hg38). For sample-wise quality control, individuals exhibiting a discrepancy between genetically inferred sex and reported sex in registries, high genotype missingness (>5%), and excess heterozygosity (±4 standard deviations) were excluded. For variant-level QC, variants with high missingness (>2%), low Hardy–Weinberg equilibrium (PP<P1P×P10^-6^), and a minor allele countP<P3 were filtered out. Chip-genotyped samples were pre-phased with Eagle 2.3.5 and imputed with the Finnish-specific SISu v4.2 imputation reference panel using Beagle v4.1 software (beagle.27Jan18.7e1.jar). Post-imputation quality control involved excluding variants with INFO score <0.7^43^.

All EstBB participants have been genotyped at the core genotyping lab of the Institute of Genomics, University of Tartu, using Illumina Global Screening Array v3.0_EST. Samples were genotyped, and PLINK format files were created using Illumina GenomeStudio v2.0.4. Individuals were excluded from the analysis if their call rate was <95%, if they were outliers of the absolute value of heterozygosity (>3 SD from the mean) or if the sex defined based on heterozygosity of the X chromosome did not match the sex in phenotype data^46^. Before imputation, variants were filtered by call rate <95%, HWE PP<P1P×P10^−4^ (autosomal variants only), and minor allele frequency <1%. Genotyped variant positions were in build 37 and were lifted over to build 38 using Picard. Phasing was performed using the Beagle v5.4 software^47^. Imputation was performed with Beagle v5.4 software (beagle.22Jul22.46e.jar) and default settings. The dataset was split into batches of 5,000. A population-specific reference panel consisting of 2,695 WGS samples was utilized for imputation, and standard Beagle hg38 recombination maps were used. Based on the principal component analysis, samples which were not from European ancestry individuals were removed. Duplicate and monozygous twin detection was performed with KING 2.2.7, and one sample was removed from the pair of duplicates^48^.

### GWAS

GWAS in FinnGen was conducted with REGENIE v3.3^49^ using the FinnGen pipeline for binary traits (https://github.com/FINNGEN/regenie-pipelines), adjusting for age, sex, genotyping chip, batch and the first ten genetic principal components.

Association analysis in the Estonian Biobank was carried out for all variants with an INFO score > 0.6 using the additive model as implemented in REGENIE v4.0 with standard binary trait settings. Logistic regression was carried out with adjustment for current age, age², sex and ten PCs as covariates, analyzing only variants with a minimum minor allele count of 10.

For the UK Biobank, MVP and All of Us, we used publicly available summary statistics of European ancestry for herpes zoster, as described in the Cohorts section. Association analyses in these cohorts were performed by the original studies (UK Biobank^50^, MVP^51^ and All of Us^52^).

### Sensitivity analyses in FinnGen

To assess the influence of control selection and immunosuppression, we ran three additional GWASes in FinnGen with REGENIE using the same settings as in the main analysis. First, controls were restricted to individuals aged at least 70 years at death or end of follow-up. Second, controls aged at least 70 years who had received a zoster vaccination in the FinnGen vaccination register were additionally excluded. Third, cases and controls with severe immunocompromise recorded at any time were excluded: HIV infection (ICD-10 B20–B24, Z21), hematological malignancy (C81–C96), primary immunodeficiency (D80–D84) or organ or bone marrow transplantation (Z94.0–Z94.4, Z94.8, T86) in hospital or drug reimbursement registers, or purchases of immunosuppressants (ATC L04) or antineoplastic agents (L01, excluding topical fluorouracil, L01BC02). We compared the effects of the eight COJO lead variants with the main FinnGen analysis using the slope of a regression through the origin and the Pearson correlation.

### Harmonization and meta-analysis

Before meta-analysis, the summary statistics of each cohort were harmonized to the GRCh38 reference genome. The genome build of each file was verified to be GRCh38 and alleles were aligned to the reference genome, strand flips were corrected, and ambiguous palindromic variants (A/T and C/G) were removed unless their strand could be resolved from allele frequencies. Effect sizes were expressed as log odds ratios (beta) for the alternative allele, and each variant was identified by chromosome, position, reference and alternative allele (SNPID, chrom:pos:ref:alt). Variants with minor allele frequency < 0.001 or imputation INFO < 0.3 were excluded. The quality of each harmonized file was checked with QQ plots and genomic inflation factors, by comparing reported P-values with P-values computed from beta and standard error, and by comparing allele frequencies with gnomAD (Supplementary Figures 1–5).

We performed a fixed-effects inverse-variance weighted meta-analysis with METAL^53^ (SCHEME STDERR), using SNPID as the marker and no genomic control correction. Heterogeneity between cohorts was assessed with Cochran’s Q and the I^2^ statistic. Variants with minor allele frequency below 1% and variants present in only one cohort were excluded. The meta-analysis output was harmonized again against the reference genome, and rsIDs were added from dbSNP (GRCh38). Inflation in the meta-analysis was assessed with the genomic inflation factor (λ = 1.10) and the LD score regression intercept (1.03, ratio 0.23), indicating that the inflation was mainly due to polygenicity (Supplementary Figure 6). Genome-wide significance was set at P < 5 × 10^−8^. For Mendelian randomization, we repeated the meta-analysis without FinnGen (four cohorts, 42,398 cases and 1,174,320 controls). Throughout, odds ratios are reported with 95% confidence intervals in square brackets.

### Definition of loci and independent signals

Lead variants were identified by LD clumping in PLINK v1.9.0-b.7.11^54^ of autosomal variants with minor allele frequency (MAF) ≥ 0.01 (index P < 5 × 10^−8^, secondary P < 1 × 10^−5^, r^2^ < 0.1, window 1,000 kb). The LD reference was the European samples of the 1000 Genomes Project Phase 3 (n = 503) in GRCh38^55^, to which variants were matched by position and alleles. The same reference was used for COJO and SuSiE. A locus was defined as ±500 kb around each lead variant, overlapping loci (lead variants less than 1 Mb apart) were merged, and the variant with the lowest P-value was reported as the locus lead variant. Because the long-range LD in the major histocompatibility complex (MHC, chr6:25–34 Mb) cannot be resolved with a reference panel of this size, all lead variants in this region were collapsed into the one with the lowest P-value, and the region was studied with HLA fine-mapping instead of COJO and SuSiE.

To identify conditionally independent signals, we performed stepwise conditional and joint analysis with GCTA-COJO v1.94.1^56,57^ in each locus outside the MHC. The input was the meta-analysis summary statistics for variants within the locus boundaries with MAF ≥ 0.01 that were present in the LD reference, with the total sample size (N = 1,716,473) for every variant. Signals were selected at P < 5 × 10^−8^ with a collinearity threshold of 0.9, and variants whose allele frequency differed from the LD reference by more than 0.2 were excluded.

### SuSiE fine-mapping

Loci outside the MHC were fine-mapped with the Sum of Single Effects (SuSiE) model for summary statistics (susieR v0.14.2^58,59^) in the European LD blocks of MacDonald et al. (2022)^60^ that contained at least one variant with P < 5 × 10⁻⁸. LD between variants with MAF ≥ 0.01 was computed from the European 1000 Genomes reference with PLINK2 v2.0.0-b.1^54^. We allowed up to ten causal variants per block and used the effective sample size summed over cohorts (267,798). For each signal, the credible set was the smallest set of variants with a 95% probability of containing the causal variant. Agreement between the summary statistics and the reference LD was checked in each block with the λ parameter and outlier detection in susieR^59^ (Supplementary Table S4).

### HLA fine-mapping

The HLA region is characterized by high genetic diversity and extensive linkage disequilibrium (LD), making it challenging to identify causal variants from GWAS signals alone. To identify specific HLA alleles independently associated with shingles susceptibility, we performed HLA fine-mapping in FinnGen (data release 14) using individual-level imputed HLA allele data. Classical HLA alleles were imputed at two-field resolution with HIBAG using a Finnish reference panel^61^. Allele dosages were extracted with PLINK2.

We included HLA alleles with imputation INFO > 0.5 and allele frequency ≥ 0.5% (98 of 135 alleles). The association between each HLA allele and shingles was tested with logistic regression, adjusting for age at death or end of follow-up, sex and the first ten genetic principal components. Alleles with P < 5.1 × 10^−4^ (Bonferroni correction for 98 alleles) were considered associated. To identify independently associated alleles, we performed stepwise conditional analysis, adding the most strongly associated allele as a covariate in each round until no further allele reached nominal P < 0.05. Alleles that reached the Bonferroni threshold in their round were considered independent signals.

To test whether B*44:02 and B*44:27 represent the same signal, we fitted two models. First, to test whether the association extended to other B*44 alleles, we added the summed dosage of all B*44 alleles to a model with B*44:02 and B*44:27. With both alleles in the model, this term measures the effect of the remaining B*44 alleles. Second, to test whether the two alleles had different effects, we fitted their combined dosage together with B*44:27 and the other independent signals, so that the B*44:27 term measured the difference between the two alleles. As their effects did not differ and the other B*44 alleles were not associated, the two alleles were combined into one signal. All independent signals were then fitted together in a joint model with the same covariates. LD between the lead alleles was estimated from dosage correlations.

HLA imputation of the EstBB genotype data was performed using the SNP2HLA tool^62^. The imputation was done for genotype data generated on the Global Screening Arrays v1, v2 and v3. We performed additive logistic regression analysis with the imputed HLA alleles using REGENIE v4.0 with standard binary trait settings. Regression was carried out with adjustment for current age, age², year of birth, sex and five PCs as covariates, using ICD-10-based phenotype data from EstBB 2026v02 freeze. Analyses were performed for all HLA alleles with minor allele frequency ≥ 1% and imputation quality info score ≥ 0.9.

### Functional annotation

Each lead variant was annotated with the nearest protein-coding gene and with the most severe consequence on a protein-coding transcript using the Ensembl Variant Effect Predictor (VEP) REST API (Ensembl release 116)^63^. To identify protein-altering variants, we also annotated all variants in LD with the COJO signals and clump leads (r^2^ ≥ 0.8 within ±500 kb in the European LD reference) with VEP, including SIFT and PolyPhen predictions.

To link the loci to target genes, we looked up variants in the GTEx v10 single-tissue eQTL results through the GTEx Portal API (https://gtexportal.org) and aligned the eQTL effects to the meta-analysis effect allele (Supplementary Table S8). We included the COJO signals, the clump lead variants, and SuSiE credible-set variants with P < 1 × 10⁻⁵ that had a posterior inclusion probability ≥ 0.1 or belonged to a credible set of at most ten variants. This lookup was descriptive, and shared causal variants were tested by colocalization (see below). We also used enhancer-to-gene predictions from ENCODE-rE2G^64^ available in the Open Targets Platform^65^, queried through its GraphQL API in November 2026, for the COJO signals, the clump lead variants and their LD proxies (r^2^ ≥ 0.8 within ±500 kb in 1000 Genomes EUR, PLINK v1.9). Open Targets provides ENCODE-rE2G predictions with a score ≥ 0.6 (Supplementary Table S10).

### Colocalization analysis

To determine whether GWAS association signals share a causal variant with expression quantitative trait loci (eQTLs), we performed colocalization analysis with the coloc.abf function of the coloc R package (v5.2.3)^66^. This approach calculates posterior probabilities for five hypotheses: no association in either dataset (H0), association only in the GWAS (H1), association only in the eQTL dataset (H2), association in both datasets driven by different causal variants (H3), or association in both datasets driven by a shared causal variant (H4). We used prior probabilities p1 = p2 = 10^−4^ and p12 = 5 × 10^−6^, as recommended by Wallace^67^. A posterior probability for H4 (PP.H4) ≥ 0.8 was used as the threshold for concluding colocalization.

We tested the seven independent signals outside the MHC region identified with COJO, as the MHC region was studied with HLA fine-mapping. The GWAS input was the European five-cohort meta-analysis (70,216 cases and 1,646,257 controls), treated as a case-control trait, and eQTL data were treated as quantitative traits. For each signal, all genes with a transcription start site within ±250 kb of the signal were tested, without filtering on eQTL significance, using variants shared between the GWAS and eQTL data in the same window. Tests with fewer than 50 shared variants were excluded.

eQTL summary statistics were obtained from the eQTL Catalogue^68^ (accessed October 2026). We included 194 datasets from immune cells, whole blood, spleen and EBV-transformed lymphocytes (LCL), including stimulated and infected cell states, and from sun-exposed and non-exposed skin and tibial nerve, because VZV reactivates in sensory ganglia and causes skin lesions. GTEx v8 tissues were included through the eQTL Catalogue. For each gene, we report the dataset with the highest PP.H4 among gene–dataset pairs with an eQTL P < 10^−4^ in the region, and all tests are reported in Supplementary Table S9.

### Gene-based, gene-set and tissue expression analyses

We performed gene-based, gene-set and tissue expression analyses with MAGMA v1.08^69^ in FUMA v1.8.2^70^ using the European five-cohort meta-analysis (N = 1,716,473). Variants were mapped to protein-coding genes (Ensembl v102) within 10 kb of the gene boundaries, and gene-level P-values were computed with the European samples of the 1000 Genomes Project Phase 3 as the LD reference. The extended MHC region (chr6:25–34 Mb) was excluded, and as a sensitivity analysis we repeated the analyses including it. Gene-based P-values were Bonferroni-corrected for the number of genes tested (19,714). Gene-set analysis was performed for 17,009 curated gene sets and Gene Ontology terms from MSigDB v2023.1.Hs, and tissue expression analysis for GTEx v8 expression in 30 general and 54 specific tissues, conditioned on the average expression across tissues. P-values were Bonferroni-corrected for the number of gene sets or tissues tested.

### Tissue and cell type-specific analyses

To identify tissues and cell types relevant for shingles susceptibility, we performed stratified LD score regression (sLDSC) with the summary statistics of the European meta-analysis. sLDSC tests whether SNP heritability is enriched in a genomic annotation, conditional on a baseline model of functional annotations^71^. In the tissue-group analysis, each of the ten tissue groups of Finucane et al.^71^, defined from cell-type-specific histone marks, was added separately to the baseline model (Supplementary Table S11). Significance was assessed with the one-sided P-value of the tissue-group coefficient, which tests for enrichment beyond the baseline annotations, with Bonferroni correction for ten tests. In the multitissue analysis (LDSC-SEG)^72^, each annotation was added separately to baseline model v1.1. We tested 489 chromatin-based annotations from the Roadmap Epigenomics and ENCODE projects, defined from peaks of six epigenetic marks (H3K27ac, H3K4me1, H3K4me3, H3K9ac, H3K36me3 and DNase hypersensitivity)^73,74^, and 205 gene expression-based annotations from GTEx and the Franke lab^75^. P-values of the annotation coefficients were corrected with FDR, and annotations with P_FDR_ < 0.05 were considered significant. Variants in the MHC region were not used in the regressions.

### Phenotype-wide association study (CodeWAS)

Diseases co-occurring with shingles were studied in FinnGen with CodeWAS in the Cohort Operations tool, using an age-and sex-matched case-control dataset of 27,818 cases and 201,350 controls. ICD-10 diagnosis codes (4,939 codes) and FinnGen endpoints recorded at any time during follow-up were compared between cases and controls with the chi-square or Fisher’s exact test. Because codes were counted over the lifetime, the associations describe co-occurrence and do not indicate temporal order. While matching on age and sex provides partial control for confounding, the analysis does not adjust for additional covariates. Codes with P < 0.05/4,939 = 1.0 × 10⁻⁵ and OR ≥ 1.2 or ≤ 1/1.2 were considered associated.

In the Estonian Biobank, a phenome-wide disease association scan was carried out for herpes zoster using ICD-10 code B02 as a binary phenotype. Associations between herpes zoster and other binary disease phenotypes were tested using logistic regression, adjusting for age at last follow-up or death, age², sex, and the first ten genetic principal components. Analyses were performed using EstBB data freeze 2023v01. Codes associated in FinnGen were considered replicated in the Estonian Biobank if the association had the same direction and P < 0.05 (or P < 0.05/5,508 for Bonferroni correction across the codes tested in EstBB).

### Heritability and genetic correlation

We estimated the SNP-based heritability of shingles with LD score regression (LDSC)^76^. Summary statistics were restricted to HapMap3 variants, and European LD scores from the 1000 Genomes Project were used as reference and regression weights. The sample size was the effective sample size summed over cohorts, and heritability was converted to the liability scale^77^ with a sample prevalence of 0.5 and a population lifetime prevalence of 30%. Genetic correlations between shingles and all FinnGen endpoints were estimated with bivariate LDSC^78^ with unconstrained intercepts in the FinnGen LDSC pipeline, using the European five-cohort meta-analysis. Further information on FinnGen endpoints is available at https://risteys.finregistry.fi/. P-values were corrected with FDR, and for plotting the endpoints were grouped by ICD-10 chapter, as for CodeWAS.

### Mendelian randomization

We tested associations between genetic liability to shingles and other diseases with bidirectional two-sample Mendelian randomization (MR) using the TwoSampleMR R package^79^. To avoid sample overlap between exposure and outcome, the shingles summary statistics came from the European meta-analysis without FinnGen (42,398 cases and 1,174,320 controls), and the other traits from FinnGen release 13 endpoints.

In the forward analysis, shingles was the exposure. Instruments were genome-wide significant variants (P < 5 × 10^−8^) clumped for independence (r^2^ < 0.001 within 10,000 kb, European 1000 Genomes reference). Because of the extensive LD and pleiotropy in the MHC region (chr6:25– 34 Mb), only the strongest instrument in this region was retained. This gave four instruments, one of which was in the MHC region. We tested twelve pre-specified outcomes selected based on earlier epidemiological evidence and the CodeWAS findings: suspected consequences of shingles (stroke, myocardial infarction, dementia, Alzheimer’s disease, multiple sclerosis, Guillain–Barré syndrome, giant cell arteritis, Parkinson’s disease and Bell’s palsy), diseases with shared or previously reported genetic susceptibility (herpes simplex infection and systemic lupus erythematosus), and fracture of the foot or ankle as a negative control outcome.

In the reverse analysis, shingles was the outcome. Instruments for each FinnGen endpoint were the lead variants from the FinnGen autoreporting (P < 5 × 10⁻⁸, one variant per locus), and endpoints with at least ten instruments were tested as exposures. The autoreporting results did not include variants in the MHC region, so no MHC instruments were used. Endpoints overlapping the shingles definition (11 endpoints: zoster, varicella, and broad viral and skin infection endpoints) were excluded, as were endpoints not defined by disease diagnoses (21 endpoints: drug purchases, procedures, anthropometric measures and smoking), leaving 334 disease endpoints (Supplementary Table S17).

Exposure and outcome alleles were harmonized with TwoSampleMR. The inverse-variance weighted (IVW) method^80^ was the primary estimator, with MR-Egger^81^, weighted median^82^ and weighted mode^83^ as sensitivity analyses. Heterogeneity was assessed with Cochran’s Q, and horizontal pleiotropy with the MR-Egger intercept (nominal P < 0.05). We also performed leave-one-out analyses and MR-PRESSO^84^, and in the forward analysis repeated the IVW analysis without the MHC instrument. P-values were corrected with FDR separately in each direction (12 tests in the forward analysis and 334 tests in the reverse analysis). In the reverse analysis, leave-one-out and MR-PRESSO were run only for exposures with P_FDR_ < 0.05.

### Observational associations with the Mendelian randomization outcomes

To compare observational and genetic evidence, we studied the twelve MR outcome traits in FinnGen with a matched case-control design. Each shingles case was matched to up to ten controls exactly on sex and birth year and by nearest neighbor on the first ten genetic principal components with the MatchIt R package^85^. The index date was the first shingles diagnosis of the case and was assigned also to its matched controls, and controls whose follow-up ended before the index date were excluded, leaving 27,724 cases and 244,315 controls. Traits were defined from ICD-10 and ICD-9 codes in the hospital inpatient, outpatient and primary care registers (Supplementary Table S15). For each trait, we defined four exposure windows relative to the index date: ever, before the index date, after the index date (new onset, excluding individuals with the trait before the index date), and 1–90 days after the index date (new onset). Diagnoses recorded on the index date were counted only in the ever window, and new onset refers to the first diagnosis recorded in the registers. Associations were tested with conditional logistic regression on the matched sets, adjusted for the first ten principal components, using the survival R package^86^. Trait–window combinations with fewer than five exposed cases or controls were not analyzed, and P-values were corrected with FDR within each window. Foot or ankle fracture was included as a negative control outcome, as in the MR analysis.

## AI use

Claude (Anthropic) was used to assist with language editing, preparation of supplementary materials, and updating of analysis code. All AI-assisted modifications were reviewed and verified by the authors, who take full responsibility for the final manuscript and analyses.

## Data availability

The summary statistics of the shingles meta-analysis generated in this study have been deposited in the GWAS Catalog (https://www.ebi.ac.uk/gwas/) under accession code GCP001766. Individual-level data from the biobanks are available only to authorized researchers because of privacy laws.

## Code availability

The analyses used the following freely available software: REGENIE v3.3 (FinnGen) and v4.0 (Estonian Biobank) (https://github.com/rgcgithub/regenie), METAL (released 2011-03-25, https://csg.sph.umich.edu/abecasis/metal/), LDSC v1.0.1 (https://github.com/bulik/ldsc), PLINK v1.9.0-b.7.11 (https://www.cog-genomics.org/plink/1.9/), PLINK2 v2.0.0-b.1 (https://www.cog-genomics.org/plink/2.0/), GCTA v1.94.1 (https://yanglab.westlake.edu.cn/software/gcta/), susieR v0.14.2 (https://github.com/stephenslab/susieR), coloc v5.2.3 (https://chr1swallace.github.io/coloc/), TwoSampleMR v0.6.7 (https://github.com/MRCIEU/TwoSampleMR), and the MatchIt v4.7.2 and survival v3.6-8 R packages (https://cran.r-project.org/).

The following web-based resources were used: FUMA v1.8.2 with MAGMA v1.08 (https://fuma.ctglab.nl/), the Ensembl Variant Effect Predictor REST API, release 116 (https://rest.ensembl.org/), the GTEx Portal API for GTEx v10 (https://gtexportal.org/), the eQTL Catalogue (https://www.ebi.ac.uk/eqtl/), the Open Targets Platform GraphQL API (https://platform.opentargets.org/), and the FinnGen Anno tool (https://anno.finngen.fi, requires a FinnGen account). CodeWAS was run in the FinnGen Cohort Operations tool. Code for the FinnGen tools is available at https://github.com/FINNGEN/.

Statistical analyses were carried out with R v4.5.3 and v4.6.0, including the packages ggplot2, RColorBrewer, locuscomparer and qqman.

## Data Availability

The summary statistics data of shingles meta-analysis generated in this study have been deposited in the GWAS catalog (https://www.ebi.ac.uk/gwas/) under accession code GCP001766. The individual-level data from the biobanks is limited to authorized researchers due to privacy laws.

## Acknowledgements

We want to acknowledge the participants and investigators of the FinnGen study.

We want to acknowledge the participants of the Estonian Biobank for their contributions. The Estonian Genome Center analyses were partially carried out in the High Performance Computing Center, University of Tartu. The Estonian Biobank Research Team was responsible for data collection, genotyping, quality control and imputation.

The authors wish to acknowledge CSC – IT Center for Science, Finland, for computational resources.

## Author contributions

H.H., H.M.O., S.S., and T.S. planned the study. H.H. and E.A. performed the analyses. H.M.O. supervised the study. The Estonian Biobank Research Team and FinnGen contributed to sample collection, genotyping, and phenotype data used in this study. H.H. wrote the original draft of the manuscript and all authors (H.M.O., S.S., T.S., A.L., V.L., M.H., S.H.) reviewed and approved the final manuscript.

## Competing interests

The authors declare no competing interests.

**Estonian Biobank Research Team**

Andres Metspalu^7^, Mait Metspalu^7^, Lili Milani^7^, Reedik Mägi^7^, Mari Nelis^7^, Tõnu Esko^7^ & Georgi Hudjashov^7^

**FinnGen**

Ville Leinonen^6^, Mikko Hiltunen^5^ & Hanna M. Ollila^1-4^

A full list of FinnGen members and their affiliations appears in Supplementary Data 1.

## Supplementary Figures

**Supplementary Figure 1.**
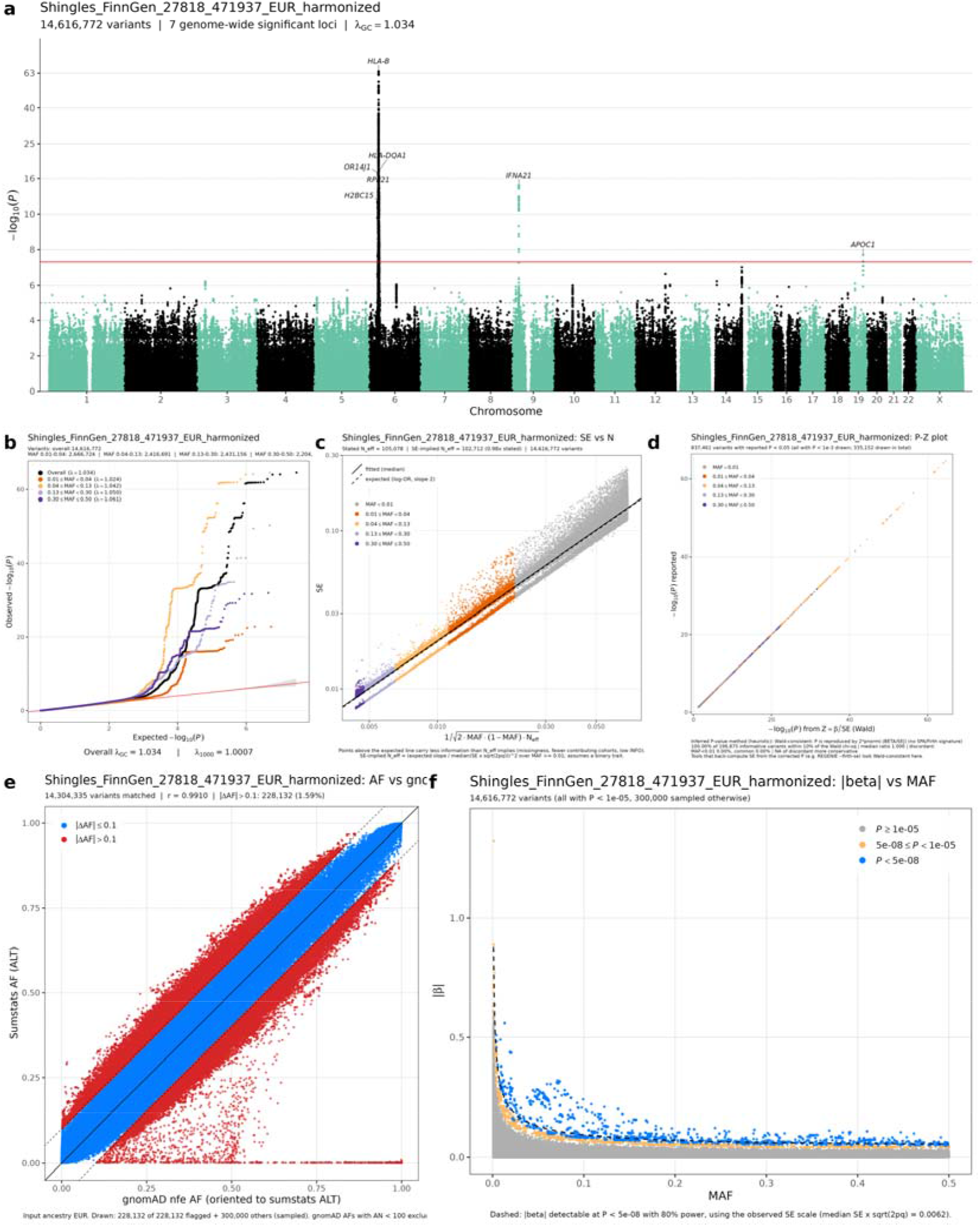
Quality control of the FinnGen summary statistics (27,818 cases and 471,937 controls, 14,616,772 variants). a, Manhattan plot. b, Quantile-quantile plot for all variants and by minor allele frequency. c, Standard error against the value expected from allele frequency and effective sample size. d, Reported P-values against P-values from the Wald statistic. e, Allele frequency against gnomAD non-Finnish Europeans. f, Absolute effect size against minor allele frequency. Most allele frequency differences from non-Finnish Europeans reflect Finnish population history.

**Supplementary Figure 2.**
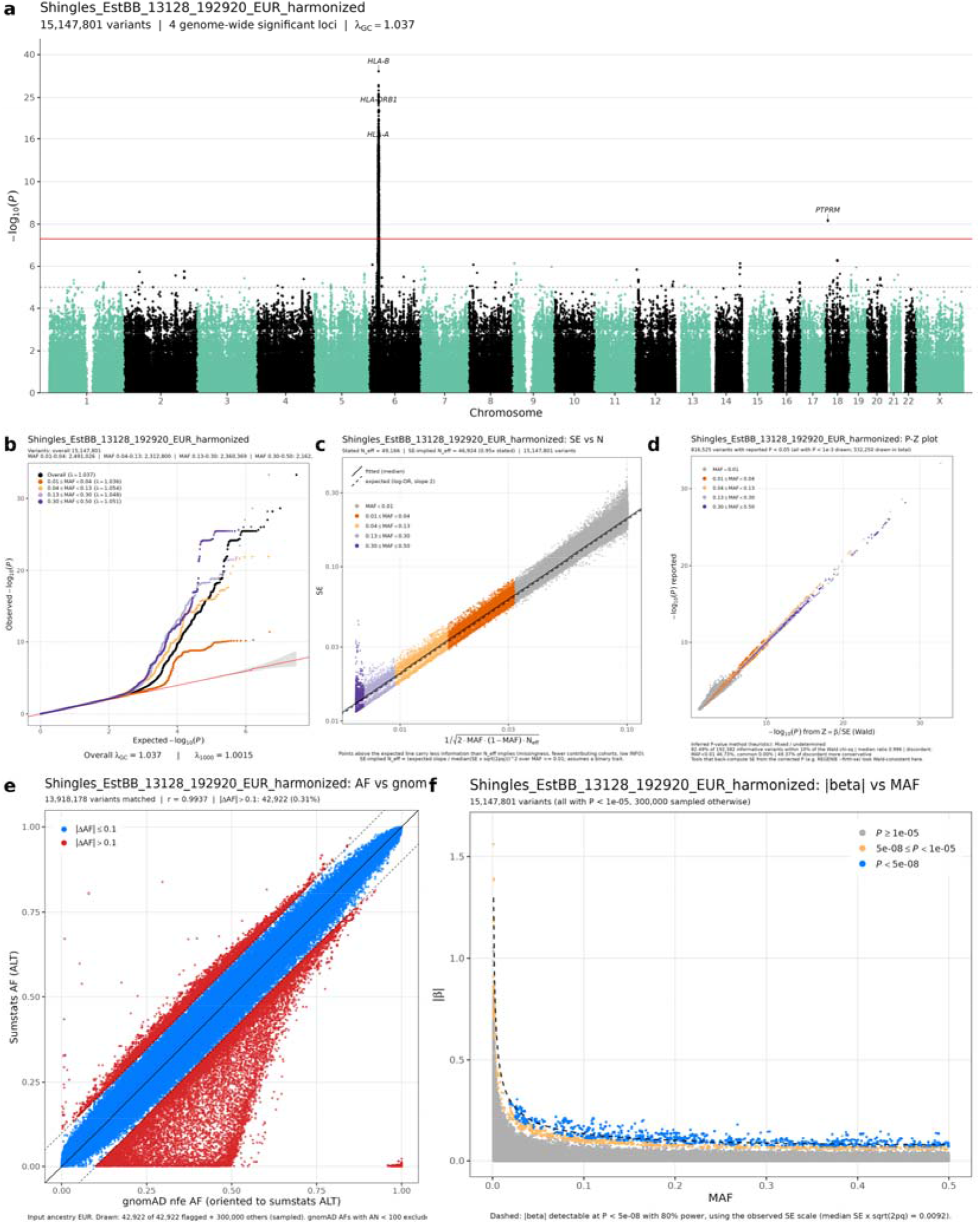
Quality control of the Estonian Biobank summary statistics (13,128 cases and 192,920 controls, 15,147,801 variants). a, Manhattan plot. b, Quantile-quantile plot for all variants and by minor allele frequency. c, Standard error against the value expected from allele frequency and effective sample size. d, Reported P-values against P-values from the Wald statistic. e, Allele frequency against gnomAD non-Finnish Europeans. f, Absolute effect size against minor allele frequency. P-values of rare variants differ from the Wald test, consistent with Firth or saddlepoint correction.

**Supplementary Figure 3.**
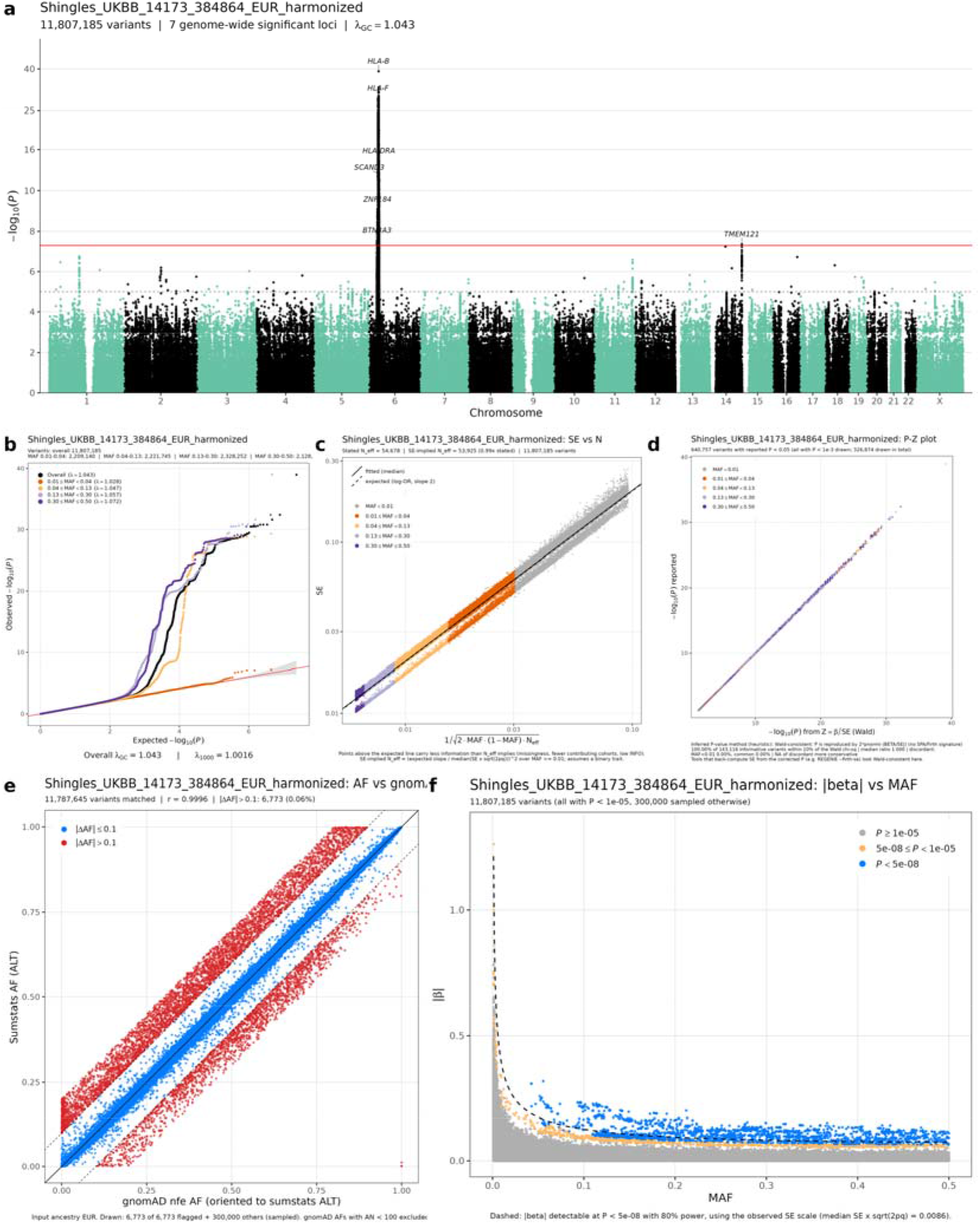
Quality control of the UK Biobank summary statistics (14,173 cases and 384,864 controls, 11,807,185 variants). a, Manhattan plot. b, Quantile-quantile plot for all variants and by minor allele frequency. c, Standard error against the value expected from allele frequency and effective sample size. d, Reported P-values against P-values from the Wald statistic. e, Allele frequency against gnomAD non-Finnish Europeans. f, Absolute effect size against minor allele frequency. Imputation quality was not available.

**Supplementary Figure 4.**
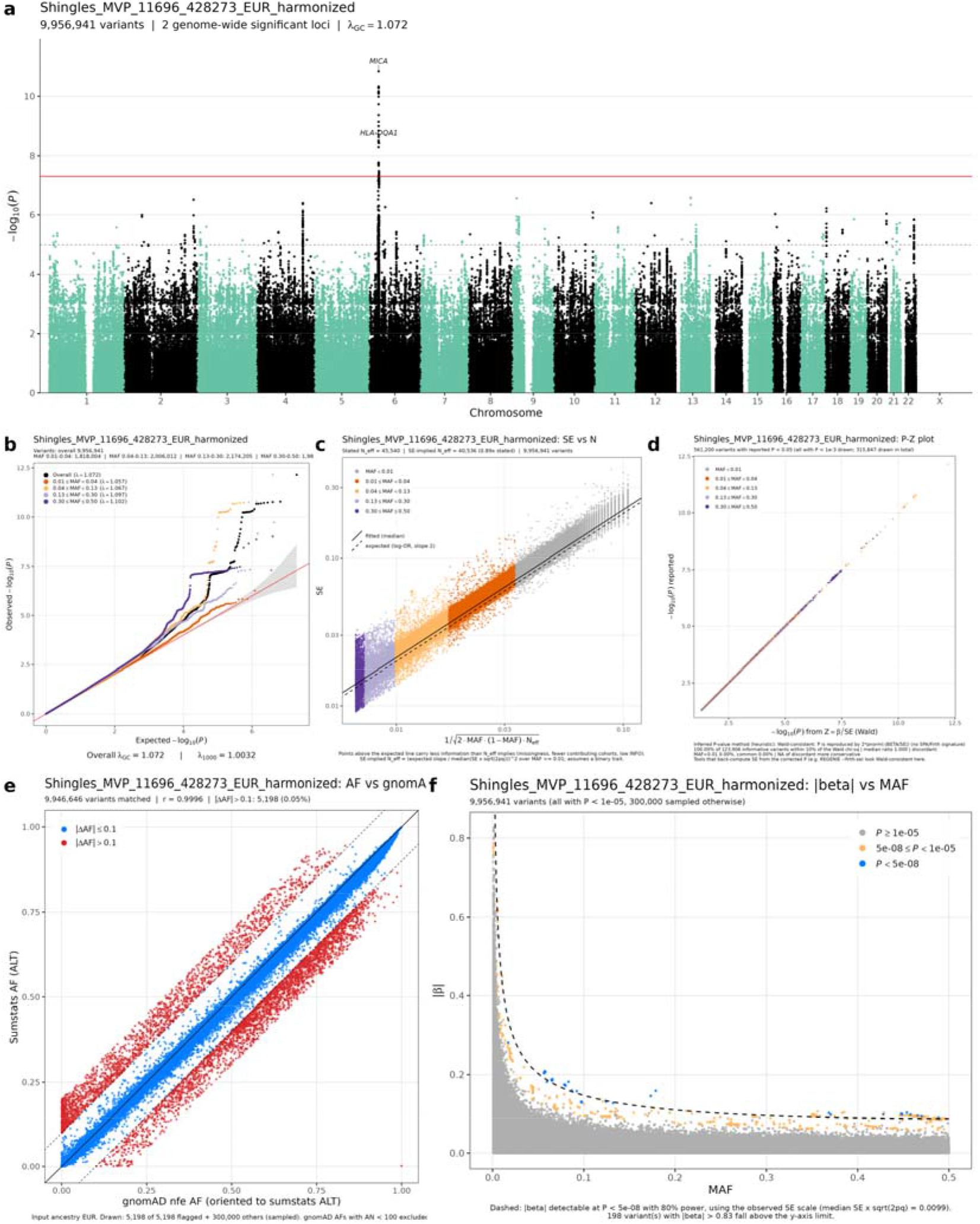
Quality control of the Million Veteran Program summary statistics (11,696 cases and 428,273 controls, 9,956,941 variants). a, Manhattan plot. b, Quantile-quantile plot for all variants and by minor allele frequency. c, Standard error against the value expected from allele frequency and effective sample size. d, Reported P-values against P-values from the Wald statistic. e, Allele frequency against gnomAD non-Finnish Europeans. f, Absolute effect size against minor allele frequency. The effective sample size implied by the standard errors is 89% of that calculated from the case and control counts.

**Supplementary Figure 5.**
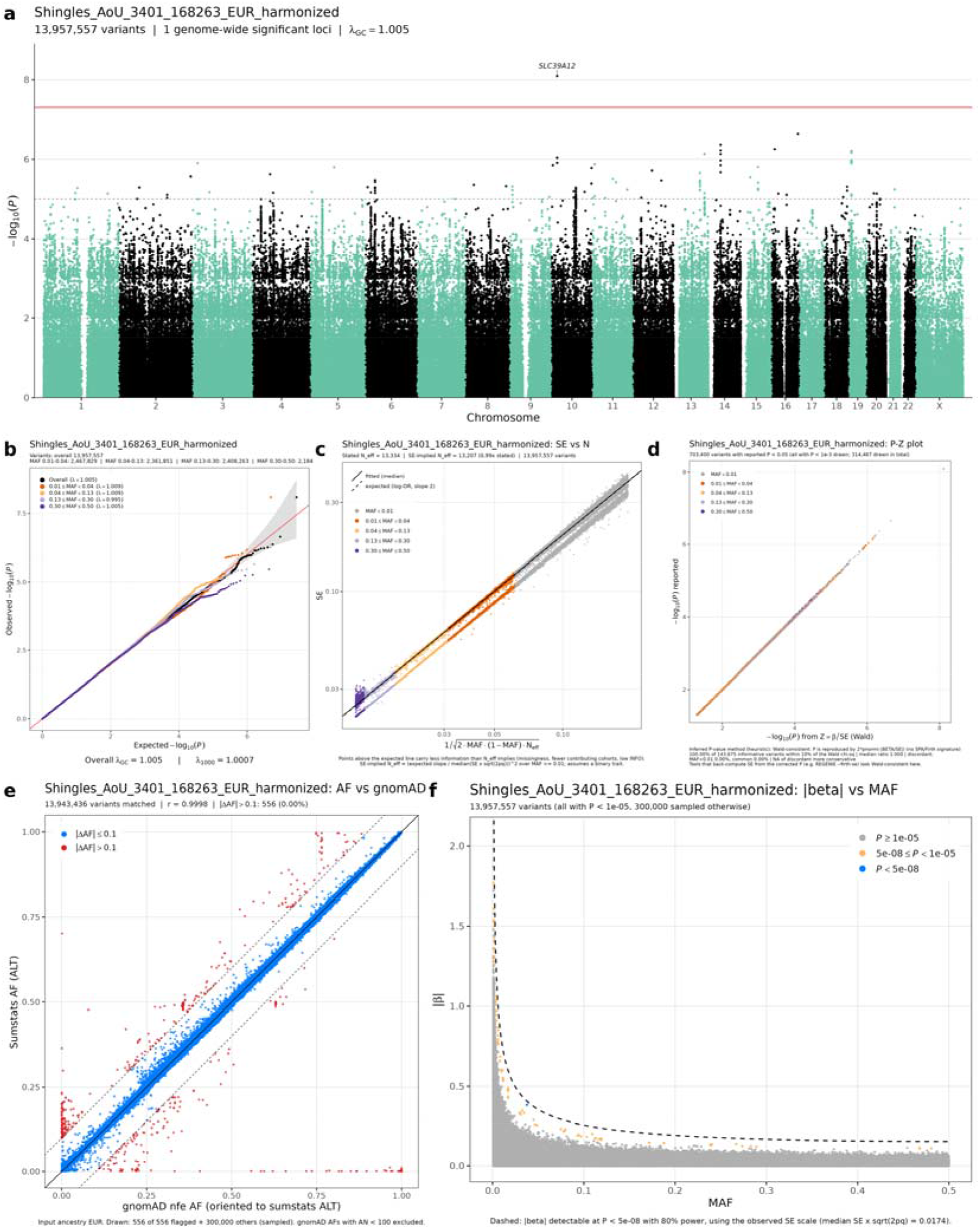
Quality control of the All of Us summary statistics (3,401 cases and 168,263 controls, 13,957,557 variants). a, Manhattan plot. b, Quantile-quantile plot for all variants and by minor allele frequency. c, Standard error against the value expected from allele frequency and effective sample size. d, Reported P-values against P-values from the Wald statistic. e, Allele frequency against gnomAD non-Finnish Europeans. f, Absolute effect size against minor allele frequency. Imputation quality was not available. The only genome-wide significant variant has a minor allele frequency of 0.04 and was not seen in the other cohorts.

**Supplementary Figure 6.**
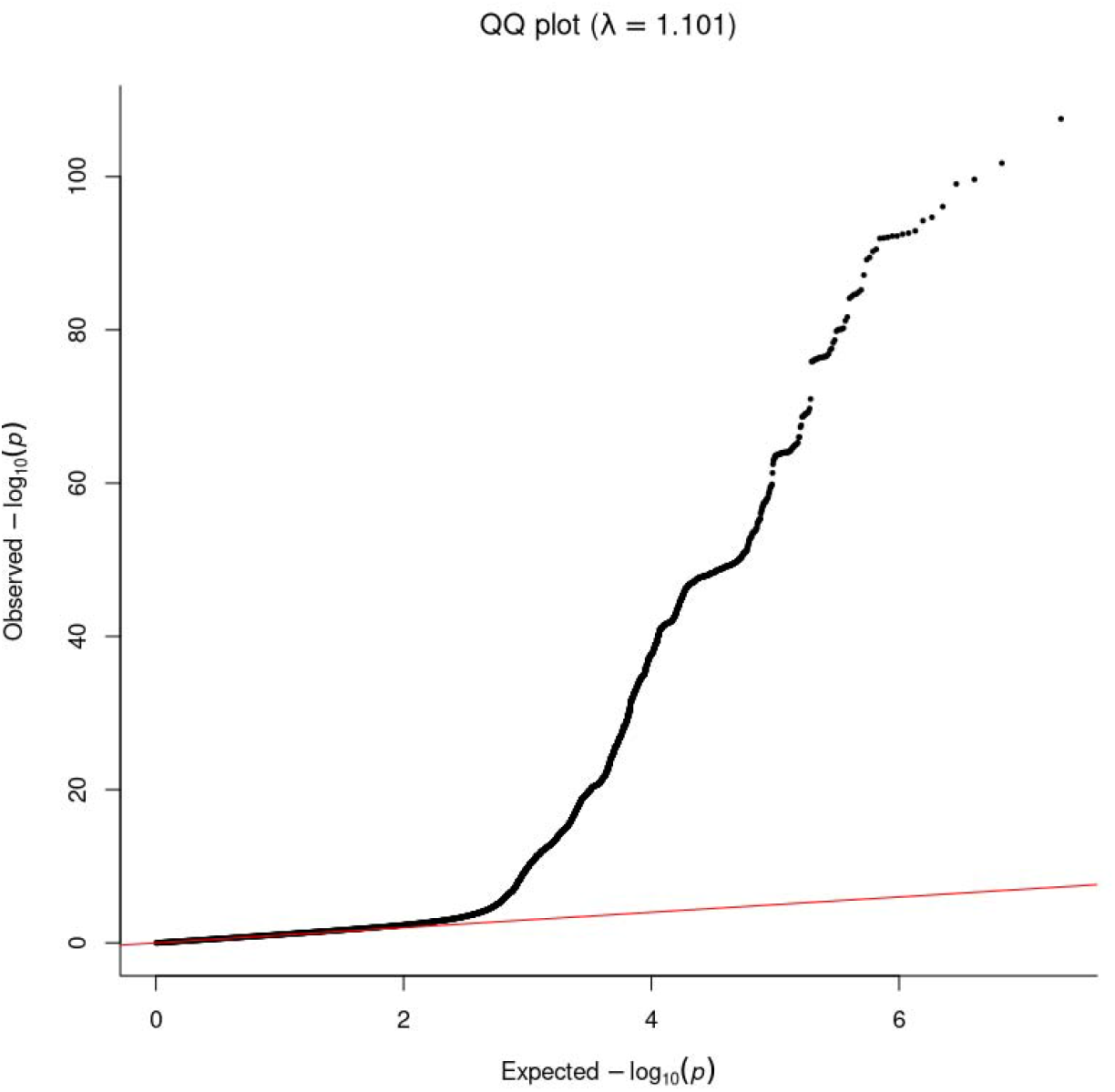
Quantile-quantile plot of the shingles meta-analysis (70,216 cases and 1,646,257 controls, 10,129,537 variants). The genomic inflation factor was 1.10, and the LD score regression intercept of 1.03 (ratio 0.23) indicates that the inflation mainly reflects polygenicity.

**Supplementary Figure 7.**
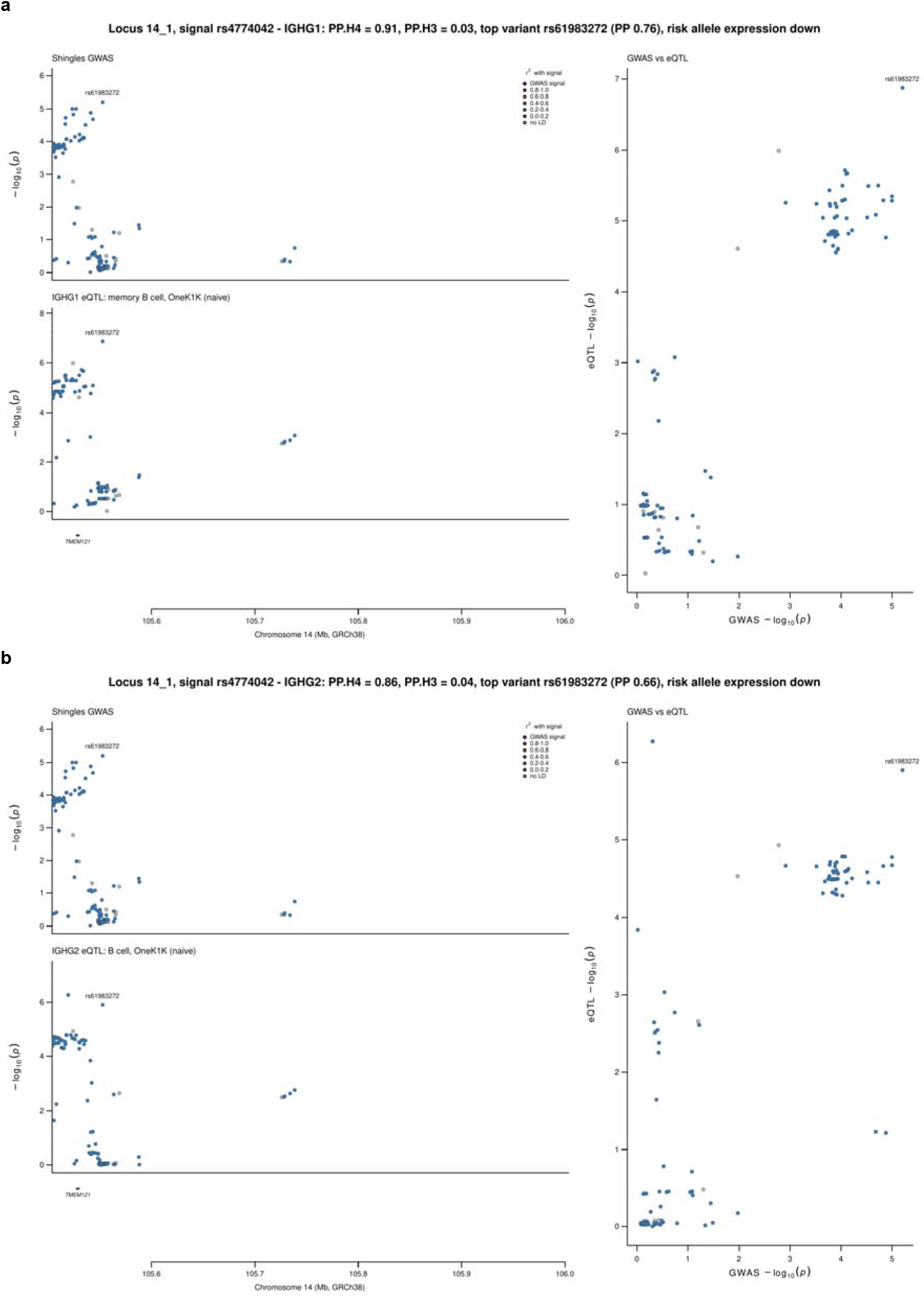
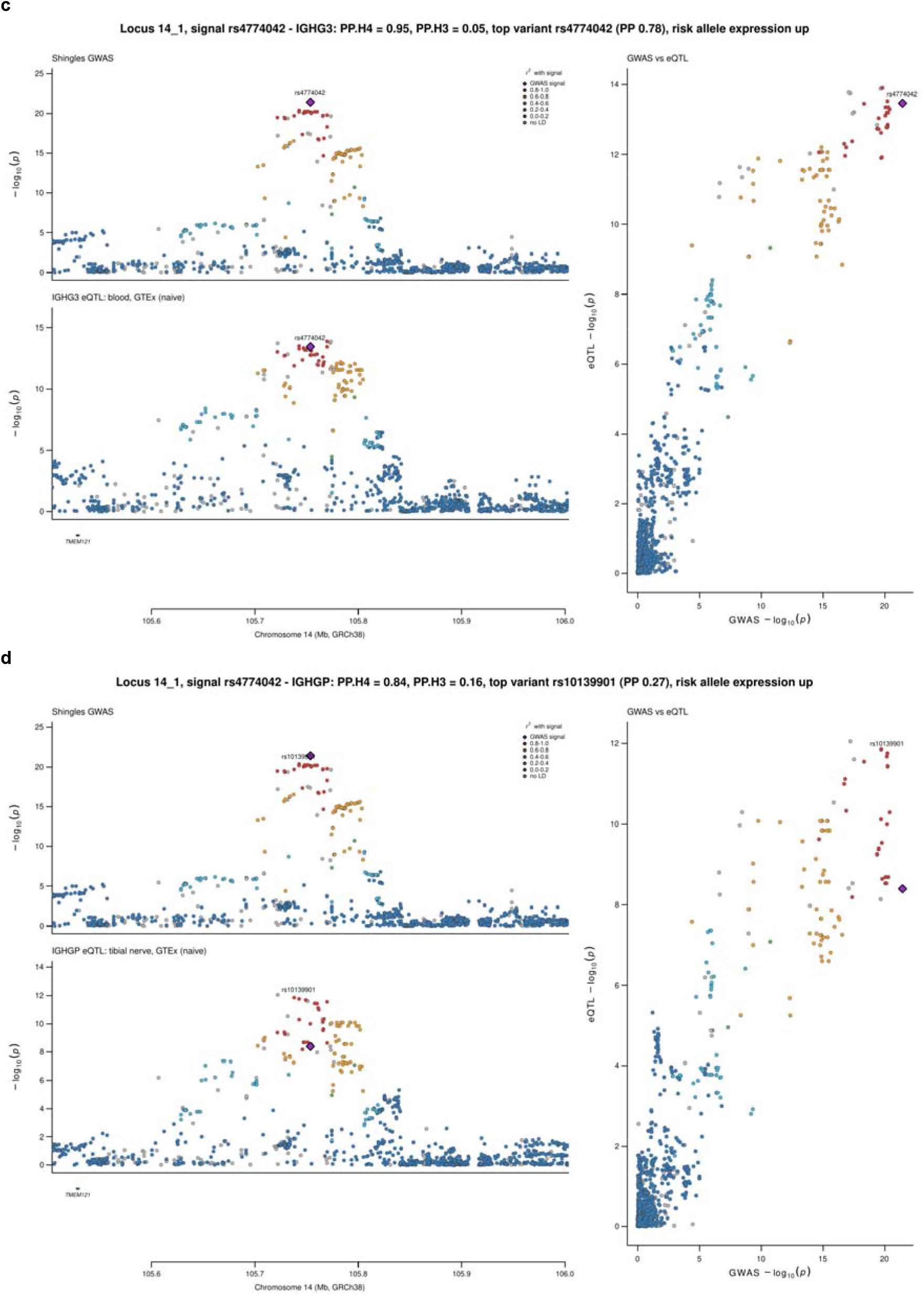
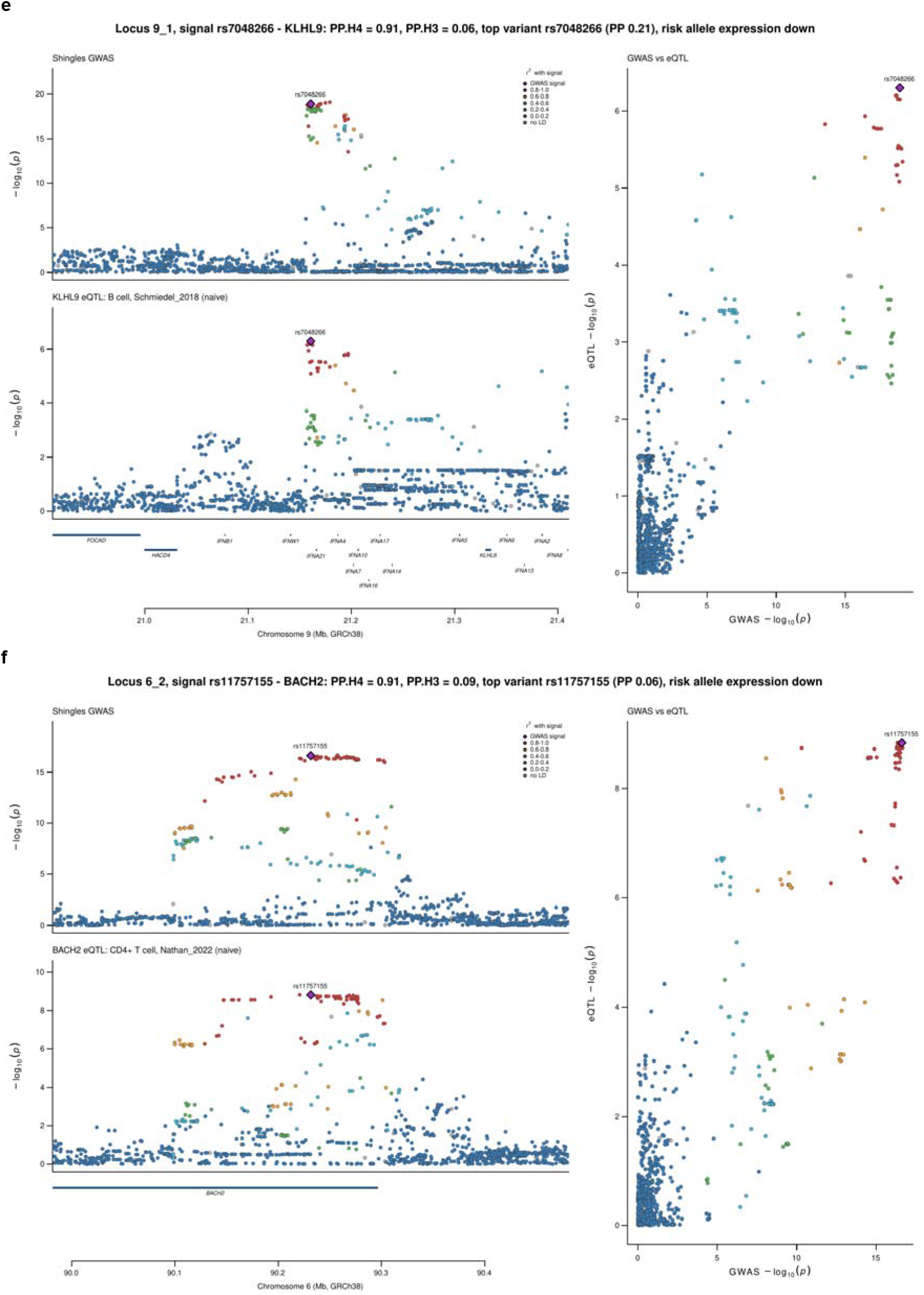
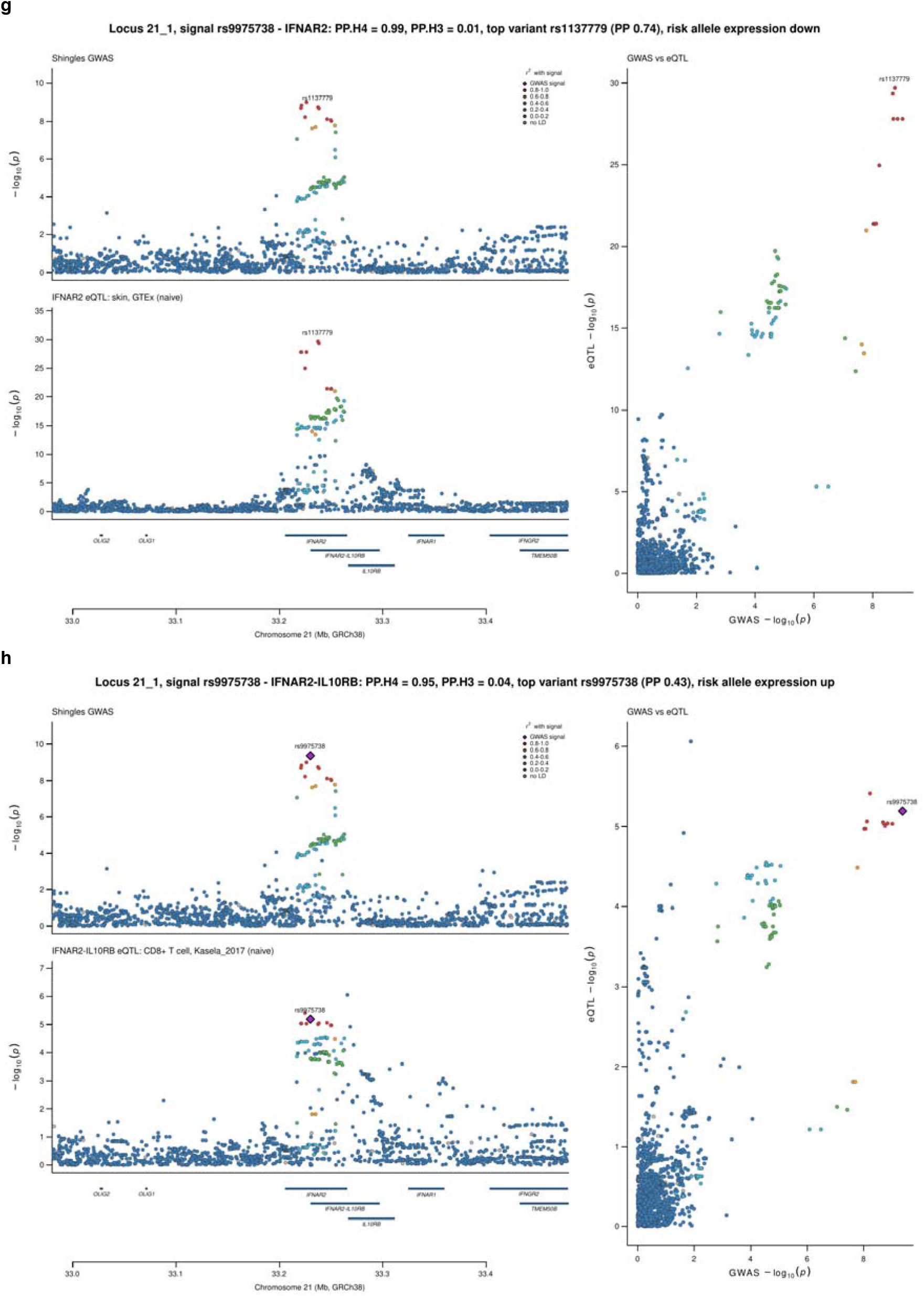
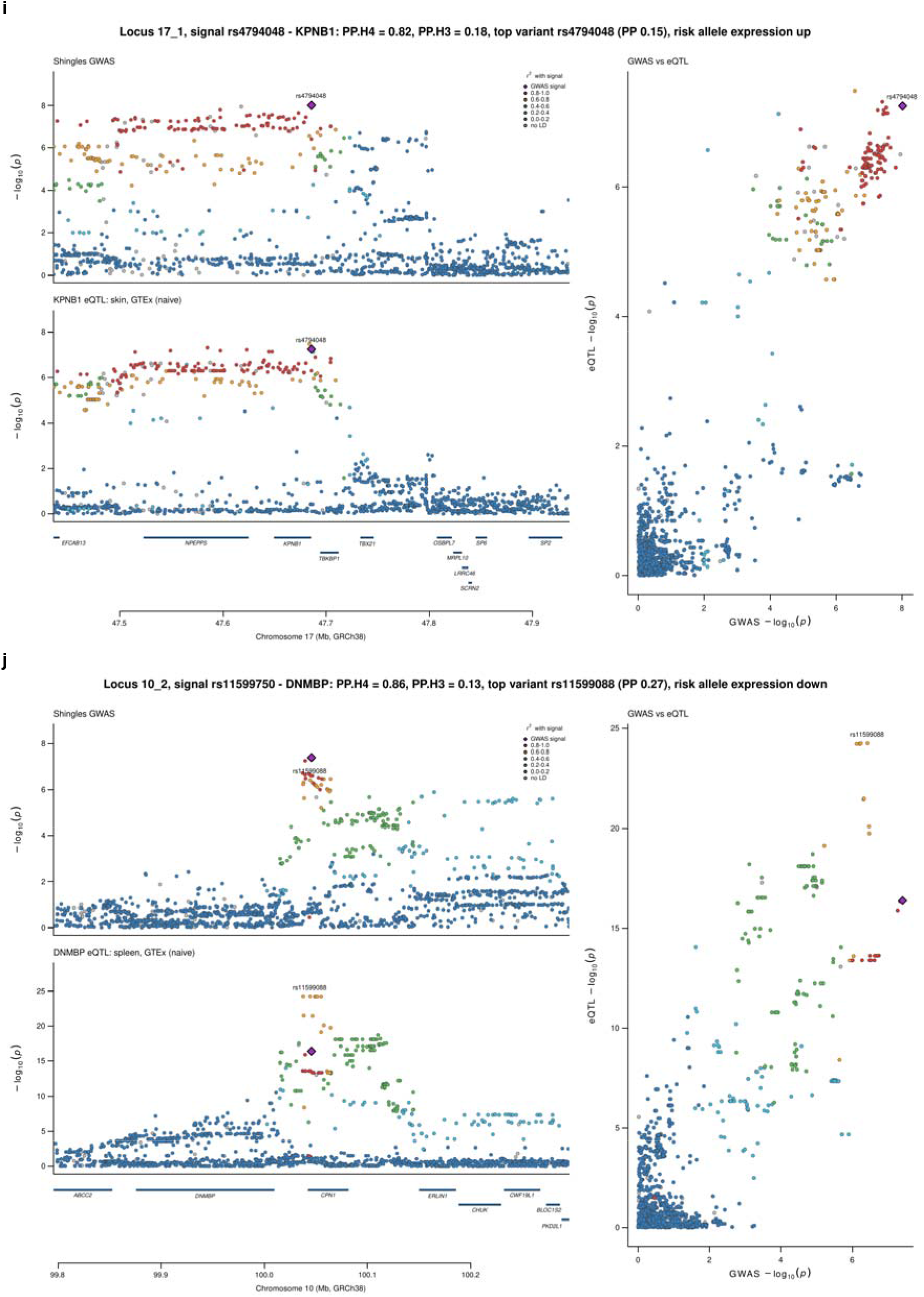
Colocalization of the shingles signals with gene expression for the genes with PP.H4 ≥ 0.8, each shown in the eQTL dataset with the highest PP.H4. a–d, *IGHG1*, *IGHG2*, *IGHG3* and *IGHGP* at the IGH locus in memory B cells, B cells, whole blood and tibial nerve. e, *KLHL9* in B cells at the *IFNA21* locus. f, *BACH2* in CD4□T cells. g, *IFNAR2* in skin. h, *IFNAR2-IL10RB* in CD8□T cells at the *IFNAR2* locus. i, *KPNB1* in skin. j, *DNMBP* in spleen at the *CPN1* locus. Left, shingles (top) and eQTL (bottom) −log_10_ P, colored by LD (r²) with the shingles lead variant. Right, shingles against eQTL −log_10_ P. PP.H4, posterior probability of a shared causal variant. All tests are in Supplementary Table S9.

